# Metformin: We need to either put it in our drinking water or rethink how we study it

**DOI:** 10.1101/2021.09.15.21263634

**Authors:** Mike Powell, Callahan Clark, Anton Alyakin, Joshua T Vogelstein, Brian Hart

**Affiliations:** Department of Biomedical Engineering, Institute for Computational Medicine, Johns Hopkins University, Baltimore, MD, USA; 2OptumLabs, Minnetonka, MN; Department of Applied Mathematics and Statistics, Johns Hopkins University, Baltimore, MD, USA; Department of Biostatistics, Johns Hopkins Bloomberg School of Public Health at Johns Hopkins, University, Baltimore, MD, USA

**Author notes:** Corresponding Author: Mike Powell, Johns Hopkins University, Department of Biomedical Engineering 3400 N. Charles St., Clark Hall 317C Baltimore, MD 21218.

**Keywords:** epidemiology, pharmacology, diabetes mellitus, retrospective cohort study, metformin

## Abstract

**Objectives:** To expose the potential impact of residual confounding in common observational study designs investigating metformin using a type 2 diabetes cohort; to propose a more robust study design for future observational studies of metformin.

**Design:** Retrospective cohort studies using a prevalent user design conducted in two distinct cohorts: individuals with type 2 diabetes and individuals with prediabetes.

**Setting:** Insurance claims database for Medicare Advantage beneficiaries in the United States, 2018-2019. An identical analysis of commercial insurance beneficiaries appears in the supplement.

**Participants:** 404,765 individuals with type 2 diabetes, 81,791 individuals with prediabetes.

**Main outcome measures:** Total inpatient admission days in 2019, total medical spend (excluding prescription drugs) in 2019. Each of these measures is treated as a binary outcome: greater than zero inpatient days and top 10% medical spend.

**Results:** We implement a common observational study design and observe a strong metformin effect estimate associated with reduced inpatient admissions and reduced medical expenditures; we also implement a more robust study design that suggests any estimated effect is attributable to residual confounding related to individuals’ overall health.

**Conclusions:** Common observational study designs examining metformin in a type 2 diabetes population are likely impacted by significant residual confounding. By additionally considering numerous negative control outcomes and a complementary prediabetes cohort, the study design proposed here demonstrates efficacy at exposing residual confounding related to overall health, nullifying the claim derived from a standard study design.

**Trial registration:** Preregistration available at https://osf.io/qf49p.

## INTRODUCTION

Metformin (a generic, glucose-lowering medication of the biguanide class) has long been a frequent candidate for repurposing as reported by a broad range of observational studies examining nondiabetes conditions and events, including postoperative mortality,[1] asthma incidence [2] and exacerbations,[3] chronic obstructive pulmonary disease-related emergency room visits and hospitalizations,[4] acute kidney injury-related ICU mortality,[5] heart failure outcomes,[6] and age-related macular degeneration.[7] Common observational study designs examining metformin compare type 2 diabetes metformin users against nonusers [1,3–6] or against insulin users;[2, 8] others pool heterogeneous groups of people with or without type 2 diabetes exposed to metformin versus no metformin.[2, 7] These and other observational studies of metformin, with varying levels of preclinical or prospective support, have suggested a host of pleiotropic benefits associated with metformin, including cancer treatment and prevention, [9, 10] anti-aging,[11] neurodegenerative disease prevention,[12] and mitigation of sepsis mortality.[13]

A principal challenge in the study of metformin using common retrospective cohort study designs is metformin’s lack of an active comparator. When used to treat type 2 diabetes, its ubiquitous use as a first-line therapy and eventual dropoff with increased disease severity means that it is prescribed in a way that a comparison to any other group (e.g., nonusers or insulin users) must address significant residual confounding concerns. The progression toward type 2 diabetes takes years, and the current care path defines two distinct states along this trajectory: prediabetes and type 2 diabetes. For individuals with prediabetes who are at high risk for progressing to type 2 diabetes, treatment with metformin is strongly recommended to prevent or delay the development of type 2 diabetes.[14] For those who receive a diagnosis of type 2 diabetes, the typical pharmaceutical treatment plan begins with metformin, may involve the addition of other agents, and often transitions to insulin as the disease progresses.[15–17] This treatment paradigm implies that metformin users generally represent the portion of the prediabetes population with the most severe disease and the portion of the type 2 diabetes population with the least severe disease. Thus, in two populations where one can expect to observe metformin use, there are known differences in disease severity between metformin users and any potential comparison group.

To assess whether the most common observational study designs are sufficiently rigorous to support so many wide-ranging metformin claims, we design a similar observational study investigating the effect of metformin in a type 2 diabetes population on two general healthcare outcomes. We then expand this common study design to include negative control outcomes (i.e., outcomes with no direct, mechanistic connection to metformin) and a *complementary cohort* (i.e., prediabetes) where we expect the bias observed in the type 2 diabetes population to be reversed. Both strategies provide valuable perspective for assessing the validity of our primary results. In the type 2 diabetes cohort, we demonstrate that metformin users have an advantage across a wide range of health outcomes, an advantage that is unattributable to metformin. We show that standard metformin study designs can produce misleading results by not sufficiently identifying and addressing residual confounding, a challenge we address by presenting a more comprehensive study design that exposes the presence and magnitude of residual confounding related to overall health in an observational metformin study.

## METHODS

### Study Design

The study presented here follows a common retrospective cohort design comparing metformin users to insulin users from a type 2 diabetes population (see the comparison of metformin users to nonusers in the supplement). We expand this basic design to include a second cohort in which we compare metformin users to nonusers from a prediabetes population, an approach we call the *complementary cohort design*.

### Data and Study Population

#### Data

This study used de-identified administrative claims data for individuals in a research database from a single, large US health insurance provider (see supplement for details on the UnitedHealth Group Clinical Discovery Database). The index date for the study was January 1, 2019. Calendar year 2018 represents the historical period of observation for all individuals in the study (i.e., pharmacy claims indicating treatment, medical claims documenting comorbidities and utilization, and other covariates are all observed in 2018). Calendar year 2019 is used to observe selected outcomes.

#### Study Population

Eligible individuals are 18-89 years old, have both medical and pharmacy coverage through a Medicare Advantage plan with 24 months of continuous enrollment spanning 2018-2019, and have at least one medical claim with a primary, secondary, or tertiary diagnosis of prediabetes or type 2 diabetes in 2018. The type 2 diabetes cohort served as the primary analysis cohort because it represents the majority of metformin users and conforms to the standard practice in the literature. Secondary analyses that consider a commercially insured population -- on average, a younger group with fewer comorbidities -- in place of the Medicare Advantage population are presented in the supplement.

Table 1 contains a summary of the available sample sizes for the type 2 diabetes and prediabetes cohorts. The analysis that follows considers metformin users versus insulin users in the Medicare Advantage type 2 diabetes population and metformin users versus nonusers in the Medicare Advantage prediabetes population, leaving all other analyses (including metformin users verses nonusers in type 2 diabetes) for the supplement. Expanded definitions providing a precise listing of qualifying generic drug names and International Classification of Diseases (ICD-10) codes, as well as other claims-based logic, appear in the supplement.

**Table 1.** Sample sizes for the various cohorts under study. Metformin and insulin users met days supply and recency requirements for 2018. Nonusers had no pharmacy claims for any diabetes drug in 2018. Excluded individuals failed to qualify for the metformin, insulin, or nonuser groups as defined in the Exposure subsection (insufficient days supply, insufficient recency, and/or prescribed both metformin and insulin at some point in 2018). Pre- and post-adjustment covariate balance for these cohorts appears in Figure 1, Figure 2, and in the supplement.

| Attribute | Commercially Insured Prediabetes | Commercially Insured Type 2 Diabetes | Medicare Advantage Prediabetes | Medicare Advantage Type 2 Diabetes |
| --- | --- | --- | --- | --- |
| Cohort Size | 17,256 | 101,826 | 83,901 | 926,977 |
| Metformin User | 1,176 | 36,998 | 3,086 | 331,085 |
| Insulin User | 0 | 6,203 | 0 | 73,679 |
| Nonuser | 14,762 | 22,447 | 78,705 | 228,934 |
| Excluded | 1,318 | 36,178 | 2,110 | 293,279 |

### Outcome, Exposure, and Covariate Definitions

#### Outcome

We show results for two outcomes: inpatient admission days and total annual medical spend (excluding prescription drugs). With these outcomes we intentionally avoid any connection to one disease or body system over another, allowing our models to rely on a standard set of baseline covariates applicable to any type 2 diabetes study. The two example primary outcomes are each thresholded: inpatient admission days (binary: >0 days) and total medical spend (binary: ≥90th percentile for the respective cohort). The cost outcome includes medical claims only due to the significantly different pharmaceutical costs between exposure groups (i.e., insulin cost > metformin cost > cost of no diabetes drugs). For each of 50 negative control outcomes (selected based on the absence of a well-established direct, mechanistic connection to metformin, e.g., toenail fungus and low back pain), the binary outcome of interest is the presence or absence of at least one claim documenting a qualifying ICD-10 code for the respective outcome in 2019.

#### Exposure

There are three exposure groups in use in the various cohorts. Nonusers are individuals who have no pharmacy claim for any diabetes drug in 2018. Metformin users are individuals with a cumulative ≥90 day supply of metformin in 2018, a daily supply of metformin lasting to at least the final 30 days of 2018, and no supply of any insulin drug in 2018. Insulin users are individuals with a cumulative ≥90 day supply of insulin in 2018, a daily supply of insulin lasting to at least the final 30 days of 2018, and no supply of metformin in 2018. Any individual who failed to meet any of these three exposure group definitions was excluded. Note that these metformin and insulin user definitions do not account for the presence or absence of any additional diabetes drug classes that might be used differentially among the exposure groups. The analysis presented in the main text compares metformin users to insulin users in the type 2 diabetes cohort, while secondary analyses that use nonusers as the comparator group are presented in the supplement. All analyses of the prediabetes cohort compare metformin users to nonusers.

#### Covariates

All adjusted models include observed covariates obtained from 2018 enrollment and medical claims data documenting individuals’ sex, age (and age squared and age cubed), urban/suburban vs. rural residence (determined by individuals’ postal codes), number of inpatient admission days, record of at least one wellness visit, presence of a medical or pharmacy claim for an influenza vaccine, Elixhauser Comorbidity Index (in-hospital mortality) score,[18] history of the outcome (i.e., at least one instance of a qualifying ICD-10 code for the same outcome in 2018), and the seven Diabetes Complications Severity Index (DCSI) [19] component scores: retinopathy (0/1/2), nephropathy (0/1/2), neuropathy (0/1), cerebrovascular (0/1/2), cardiovascular (0/1/2), peripheral vascular disease (0/1/2), and metabolic (0/1/2). Inpatient stays and Elixhauser Comorbidity Index scores are log transformed (*x*′ = *ln*(*x* + 1)).

### Statistical Analysis

The following procedure explains how we obtain odds ratios and confidence intervals for each of the two example outcomes and all of the negative control outcomes in each cohort:

1. Subset the cohort to only those individuals meeting one of the two exposure definitions (e.g., metformin users and insulin users in type 2 diabetes).
2. Conduct an unadjusted analysis using Fisher’s exact test to obtain an odds ratio between the two exposure groups and a corresponding confidence interval.
3. Estimate the propensity for metformin treatment using logistic regression and all observed covariates listed previously.
4. Trim the sample to only include individuals from the treated and control groups such that all individuals have a propensity for metformin treatment greater than or equal to the larger of the 1st percentiles of the two exposure groups’ propensity score distributions.
5. Fit an inverse propensity-weighted (IPW) logistic regression to estimate the treatment effect of metformin using all observed covariates listed previously.

### Patient and Public Involvement

Neither patients nor the public were involved in designing this study, setting its objectives and outcome measures, interpreting the study results, or presenting the study findings.

## RESULTS

### Covariate Balance Diagnostics

Figure 1 presents a diagnostic covariate balance plot showing the pre- and post-adjustment covariate balance achieved in models for the two example outcomes. Although metformin users appear healthier across numerous indicators of health, inverse propensity weighting yields acceptable covariate balance, evidenced by the standardized mean differences for all covariates having absolute values less than 0.1.

**Figure 1.**
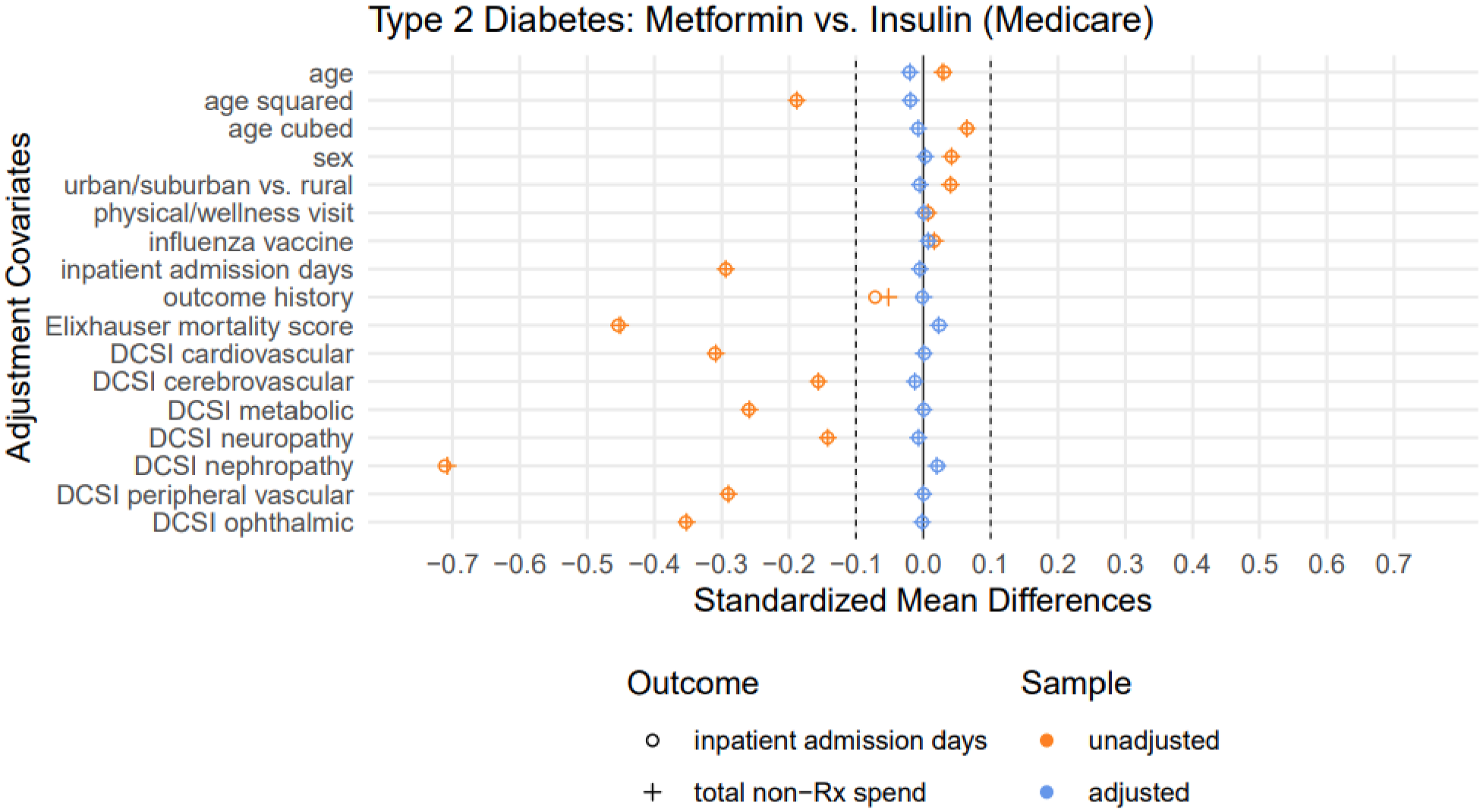
Here we show two overlaid balance plots (one for each outcome) for the type 2 diabetes cohort, each depicting both the unadjusted covariate balance and the balance achieved after adjustment using inverse propensity weighting (IPW). Negative values indicate that the metformin group has lower averages/prevalence for the indicated covariates (in the case of sex, a higher percentage of males). The unadjusted balance markers indicate that the metformin users have spent less time in the hospital, have fewer or less severe comorbidities (lower Elixhauser in-hospital mortality scores), and score significantly lower on all Diabetes Complications Severity Index (DCSI) components. These are indicators that the metformin exposure group is consistently healthier than the insulin user group. Across both outcomes, the IPW approach achieved satisfactory balance after adjusting for every observed covariate of interest.

### Primary Results

Using covariates, statistical methods, and a study design representative of what is commonly found in the literature (in this case, metformin users vs. insulin users) and demonstrating satisfactory balance on the observed covariates, we present promising results in Table 2, indicating a beneficial metformin treatment association in the Medicare Advantage type 2 diabetes population. These strong effect sizes, extremely small p-values, large E-values^1^,[20] and covariate balance plots all tempt the investigator to craft a plausible story explaining why metformin *should* have these effects.

**Table 2.**
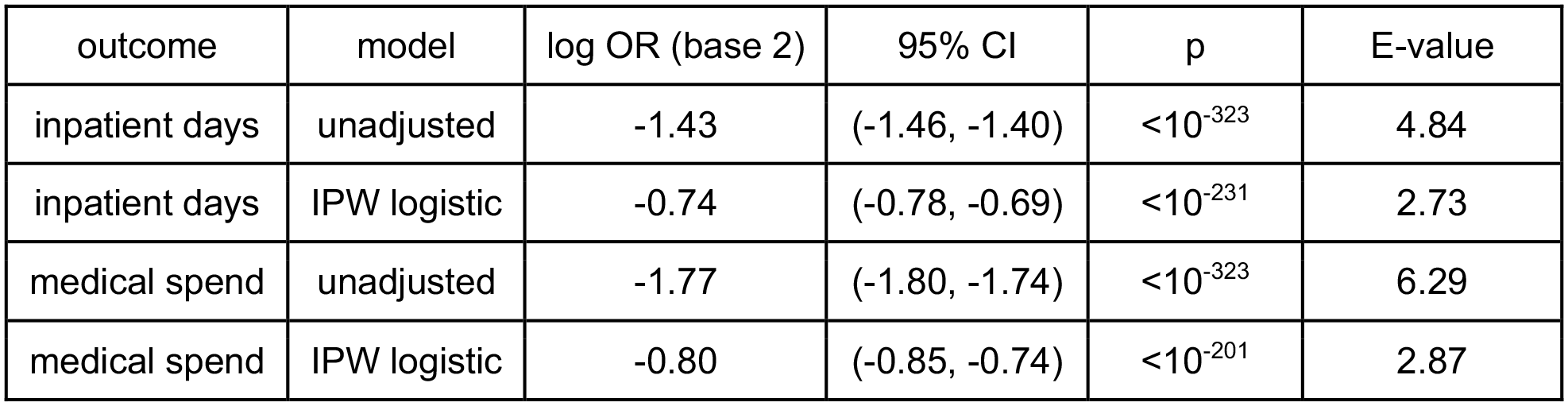
Treatment effect estimates for metformin: this study was conducted in a Medicare Advantage type 2 diabetes population comparing metformin users to a control group of insulin users. The outcomes represent >0 inpatient admission days in 2019 and a total medical spend exceeding the 90th percentile of all type 2 diabetes patient expenditures (>$25,793). With such strong estimated effect sizes and small p-values, metformin appears strongly associated with fewer inpatient admission days and lower health care costs, even after adjustment for a range of relevant covariates.

### Results in Context

Before we add another finding to the list of metformin’s many suggested benefits, we consider the same results as part of a more robust study design involving negative control outcomes and a complementary prediabetes cohort (see “Validation Tools” in the supplement for expanded discussion of how we select and implement negative controls and complementary cohorts). In short, the negative control outcomes expose the biases in the primary and complementary cohorts. Instead of focusing exclusively on the result in the primary cohort, we interpret the full results of this expanded complementary cohort design. Figure 1 showed satisfactory observed covariate balance for the type 2 diabetes cohort, while Figure 2 shows similarly satisfactory balance in the prediabetes cohort; covariate balance plots for all 50 negative control outcomes in both cohorts appear in the supplement in Figure S2.

**Figure 2.**
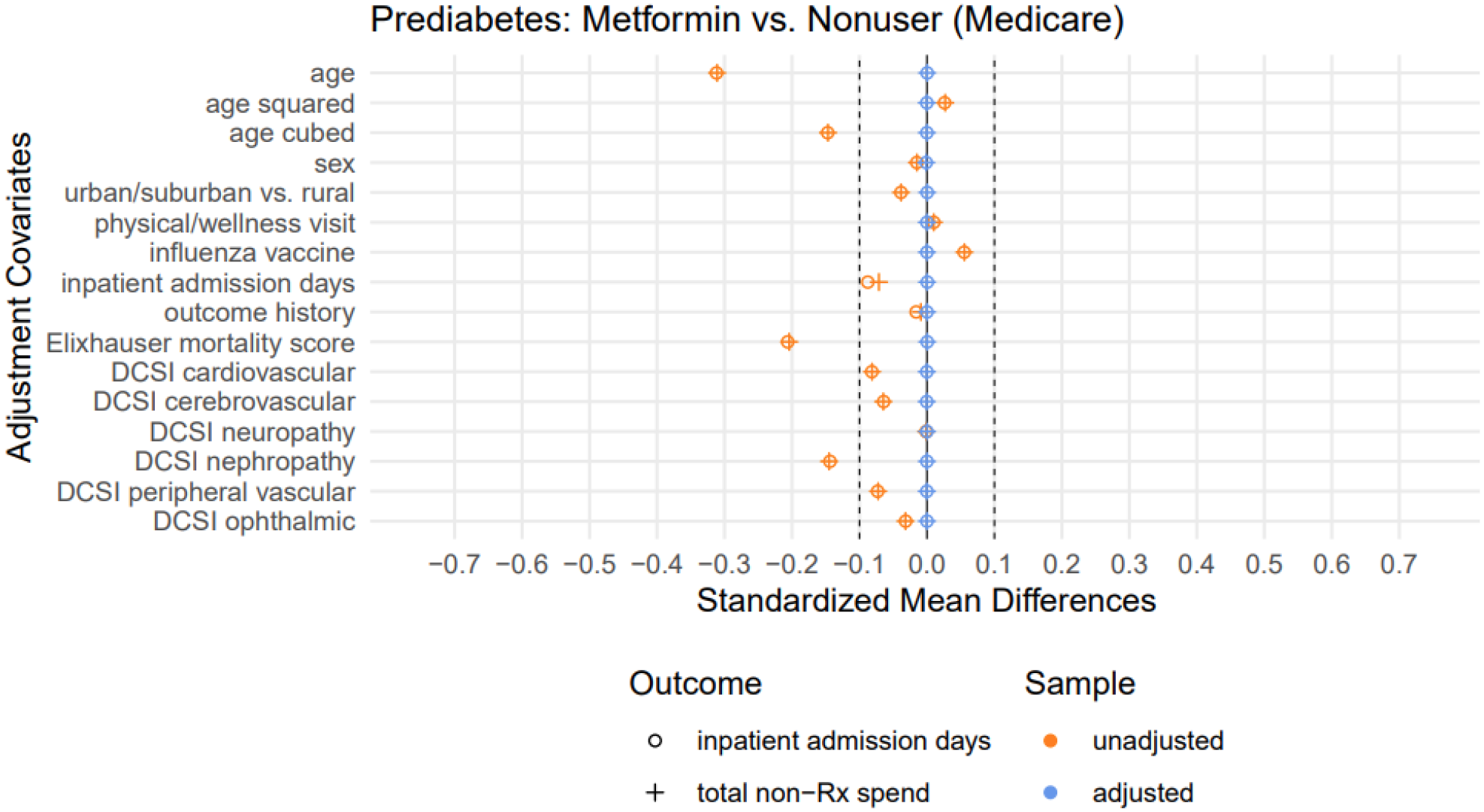
Here we show two overlaid balance plots, one for each primary outcome in the prediabetes cohort. Negative values indicate that the metformin group has lower averages/prevalence for the indicated covariates (in the case of sex, a smaller percentage of males). Across both outcomes, the IPW approach achieved satisfactory balance after adjusting for every observed covariate of interest. The observed bias related to overall health and disease severity in the unadjusted analysis is mostly nullified in comparison with the type 2 diabetes cohort depicted in Figure 1, making the prediabetes population a suitable candidate to be a complementary cohort.

Satisfied with the covariate balance achieved in these models, we now look to the forest plots in Figure 3 to examine the negative control outcome experiments in the prediabetes cohort (left) and the type 2 diabetes cohort (right). We expect negative experiments to lead to log odds ratios of zero on average. In the type 2 diabetes cohort, metformin users are biased toward lower event rates (log odds ratio <0) across a vast number of outcomes spanning many body systems. This finding suggests that, while the observed covariates in Figure 1 are well balanced after adjustment, substantial residual confounding related to overall health may be influencing our primary result. In the prediabetes cohort, the metformin users and nonusers were much more similar before balancing (see Figure 2), making the groups appear somewhat comparable. Nevertheless, this same collection of negative control outcomes exhibits the opposite bias for metformin users (i.e., a shift toward higher event rates as seen in Figure 3). Again, despite our best efforts to control for potential confounding, this result highlights the inadequacy of our initial efforts to minimize residual confounding related to overall health, and it confirms that the prediabetes cohort is biased in the opposite direction of the type 2 diabetes cohort, making it an ideal complementary cohort.

**Figure 3.**
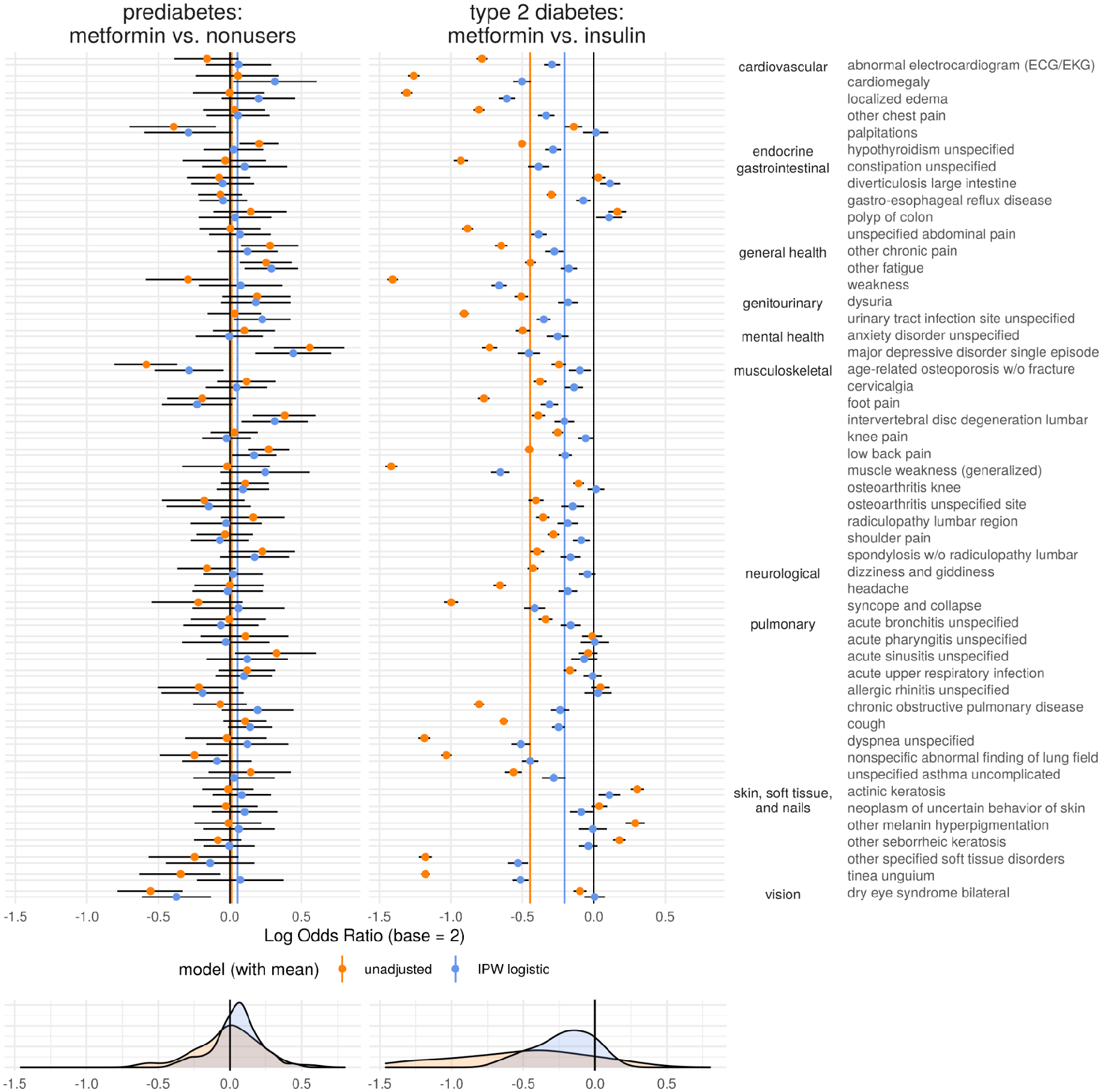
*Residual Confounding Plot:* The left forest plot depicts log odds ratios estimating the treatment effect of metformin (control group of nonusers) in 50 negative control outcome experiments in a Medicare Advantage prediabetes cohort. Average log odds ratios for the unadjusted and IPW logistic regression models are denoted by vertical orange and blue lines, respectively. While the observed covariates and analytical methods appear to produce a distribution of null results in prediabetes, there is still evidence of bias that makes metformin appear harmful (i.e., log odds ratios appear on average to be > 0). The right forest plot depicts the treatment effect estimates for metformin (control group of insulin users) in the same 50 negative control outcome experiments conducted in a type 2 diabetes cohort. The same modeling approach is more clearly insufficient in type 2 diabetes where only residual confounding can explain the multitude of significant, left-biased results in a collection of negative control outcome experiments. The outcomes listed on the far right serve as labels for all 50 experiments in each cohort and are grouped by body system to demonstrate these findings span a wide range of conditions. The bottom row depicts column-wise kernel density estimates for each distribution of point estimates. In each cohort, the density estimates for the adjusted odds ratios suggest a shift away from the null.

Finally, let us reconsider the primary outcome results in light of these negative control outcome experiments in both cohorts, all of which are depicted together in Figure 4 (Figure S3 in the supplement provides a brief tutorial on interpreting Figure 4 using various notional results). Critically, our conclusions using the combined insights from the negative control outcomes and complementary cohort design are different compared to what typical study designs would conclude. For both primary outcomes, inpatient admission days and total medical spend, we originally obtained p-values no larger than 10^-201^ for treatment effect estimates that appear to be beneficial. We even demonstrated excellent covariate balance on a reasonable set of covariates, including markers of diabetes severity. This is where many published studies stop.

**Figure 4.**
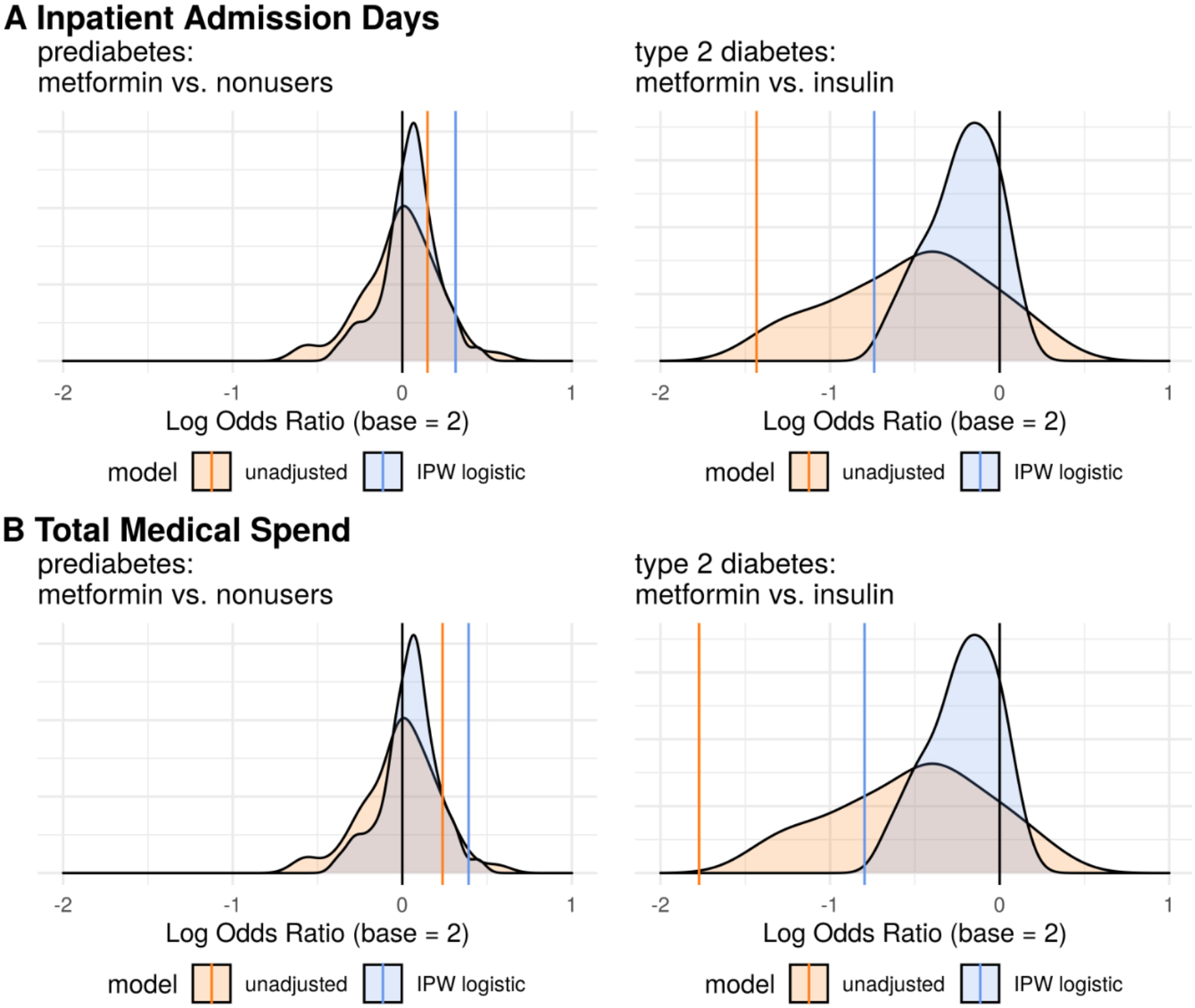
Colored vertical lines indicating the treatment effect estimates for unadjusted and adjusted models are overlaid on distributions of treatment effect estimates obtained for negative control outcomes using identical modeling approaches. For both outcomes (inpatient admission days and total medical spend), the estimated treatment effect of metformin in the type 2 diabetes population appears strong, exceeding the benefit observed for the vast majority of negative control outcomes. While that suggests the effect might be real, the effect estimates for inpatient admission days and total medical spend are both substantially reversed in the complementary prediabetes cohort. These observations suggest that there is either a heterogeneous treatment effect or significant residual confounding in the type 2 diabetes cohort, the prediabetes cohort, or both. An exploration of these possibilities is absent in much of the published observational study literature.

If we go a step further and consider a few or even 50 negative control outcomes, we might convince ourselves that the observed effect is stronger than nearly all the negative control effects (see Figure 4 type 2 diabetes subplots), indicating that while bias is clearly present, there is still hope of a real treatment effect. Proceeding to the complementary cohort, we see a result that strongly contradicts what we observe in the primary cohort. If metformin is believed to have an effect irrespective of a person’s type 2 diabetes severity, we should find similar results in the type 2 diabetes and prediabetes cohorts. Only when we take this additional step of considering the treatment in the complementary cohort do we realize the impact of health-related residual confounding on our primary result and question the validity of the finding.

## DISCUSSION

### Statement of principal findings

We show that conventional observational study designs may lead to strong results suggesting that metformin use may lead to a meaningful reduction in inpatient admissions and healthcare expenditures; we then (1) expand the conventional study design to include numerous negative control outcomes for the purpose of exposing potential bias, and (2) replicate the primary cohort analysis in a complementary cohort carefully selected to remove or reverse the bias identified in the primary cohort. Using this more comprehensive study design we find that primary results using a standard metformin study design may be unreliable and must be further validated with negative control experiments and complementary cohorts.

### Strengths and weaknesses of the study

A notable strength of this study is the deliberate inclusion of a complementary cohort (prediabetes) in which the exposed group of metformin users is expected to be less healthy than its comparison group. Instead of relying on covariate adjustments to balance the comparison groups from a position that favors the exposure, we test the ability of these same covariate adjustments to offset an expected health disadvantage. This complementary cohort approach serves a fundamentally different purpose than simply replicating the same design in different data sets where a flawed design might lead to the same issues with residual confounding. Others have described approaches similar to the study design presented here, most notably in the description of *triangulation* for epidemiology provided by Lawlor, et al.[21] We incorporate many of the same principles by generating multiple lines of evidence to evaluate a hypothesis using different populations and different control groups (e.g., multiple data sets: commercially insured vs. Medicare Advantage; multiple control groups: metformin users vs. insulin users OR metformin users vs. nonusers), conducting numerous negative control experiments, and examining the treatment effect in a complementary cohort (similar to the idea of a *cross-context* study) with presumably different biases or confounding structures.

Despite the advantages of this study design, this study suffers from other characterized limitations common to claims-based studies, such as only including individuals with qualifying health insurance, selective capture of variables associated with or incentivized by financial reimbursement, and missingness of key elements of diabetes severity such as diabetes duration.[22, 23] Additionally, the proposed design relies on a qualitative assessment of diagnostic plots presenting distributions of potentially dependent effect estimates (e.g., the negative control outcomes include related pulmonary conditions and related musculoskeletal conditions). Treating these distributions as if they represented independent events is inappropriate, and they should not be considered a valid null distribution for assessing the significance of the primary result. Also, by implying that a real effect must also be observed in the disadvantaged complementary cohort, we recognize the potential to be biased toward a type 2 error (i.e., a false negative), incorrectly rejecting a real effect that is masked by bias.

### Strengths and weaknesses in relation to other studies

Although negative control experiments are recommended as best practice by leading collaboratives such as OHDSI, negative control experiments are not commonly reported in observational drug-repurposing studies.[23] Numerous negative control experiments are even rarer. Challenges to a primary result by triangulation with a complementary cohort are virtually nonexistent. The importance of these extra validation steps stems from the inadequacy of the observable health status variables readily available in claims data to produce an unbiased estimate of the metformin treatment effect.

Some claims-based study limitations are addressed in other types of study designs, such as prospective observational studies, registry-based studies, and randomized controlled trials. Additionally, deriving baseline comorbidity profiles, medication exposure histories, or other key variables of interest can be accomplished using alternative strategies such as joining claims data to EHR data, clinical notes, or patient-reported data to obtain a more accurate view of an individual’s overall health. However, such strategies often come at the expense of sample size, and even when executed well, may still exhibit similar biases.

### Meaning of the study: possible explanations and implications for clinicians and policymakers

Despite our best efforts to eliminate residual confounding according to best practices in the metformin and observational study literature, we acknowledge and in fact demonstrate that our efforts have fallen short. This is precisely why we should collectively be concerned. We can either continue to accept the findings of conventional observational study designs at face value, or we can demand more comprehensive work be done to expose residual confounding if it exists. We believe it always exists, and in situations where there is any doubt about the comparability of various exposure groups, additional steps like those shown here must be carried out. Observational studies can motivate randomized controlled trials, influence clinical practice in settings where gold-standard prospective evidence is sparse, and get amplified by media outlets to the general public, so the cost of overvaluing a poorly designed observational study can lead to wasted research funds and even put participants’ and patients’ health at unnecessary risk.

### Unanswered questions and future research

The challenge associated with the complementary cohort design lies in the identification of a suitable complementary cohort. There are other diseases with a progressive trajectory; some have a predisease diagnosis as well as a clinical disease stage, and others are defined in multiple stages (e.g., chronic kidney disease). Applying this framework to those settings would be a natural extension; however, extending this framework to settings involving a primary cohort with one disease and a complementary cohort with a different disease would most likely introduce additional challenges.

## Data Availability

The data are proprietary and are not available for public use, but under certain circumstances, the data may be made available to approved auditors under a data use agreement to confirm the findings of the current study.

## What is already known on this topic

- Metformin has been associated with wide-ranging pleiotropic benefits in many observational studies; these studies commonly compare metformin users to nonusers or to insulin users.
- Metformin users as a group tend to represent the least healthy prediabetes individuals, yet they represent the healthiest portion of the type 2 diabetes population.
- Observational studies of metformin focus primarily on the type 2 diabetes population where metformin is widely used, but they rarely attempt to quantify or depict the degree to which residual confounding related to metformin users’ overall better health may be responsible for favorable study findings.

## What this study adds

- This study demonstrates that residual confounding is a lurking culprit among published metformin claims relying solely on observational study designs and that the more comprehensive study design executed here is capable of exposing hard-to-quantify residual confounding related to overall health.

## Contributors and Sources

Mike Powell (guarantor), Anton Alyakin, and Joshua Vogelstein are affiliated with Johns Hopkins University and have previously conducted drug-repurposing studies related to COVID-19. Callahan Clark and Brian Hart are employees of UnitedHealth Group (UHG) conducting research and development activities, often in the field of pharmacoepidemiology. Callahan Clark’s background as a pharmacist and overall familiarity with the type 2 diabetes landscape prompted the initial exploration of metformin due to the many claims in the literature that it is a widely repurposable drug. The authors from Johns Hopkins University conducted the statistical analysis of UHG claims data with assistance and oversight from the UHG authors. Several references cited in the manuscript are representative of the types of studies our example is modeled after, and the remaining references generally point to methodology papers forming the theoretical basis for our approach.

## Contributions by Author

Mike Powell (guarantor) - conceived and designed the study, analyzed and interpreted data, drafted and revised manuscript, approved final manuscript

Callahan Clark - conceived and designed the study, acquired and interpreted data, revised manuscript, approved final manuscript

Anton Alyakin - conceived and designed the study, interpreted data, revised manuscript, approved final manuscript

Joshua Vogelstein - interpreted data, revised manuscript, approved final manuscript Brian Hart - conceived and designed the study, acquired and interpreted data, revised manuscript, approved final manuscript

## Transparency Declaration

Mike Powell affirms that the manuscript is an honest, accurate, and transparent account of the study being reported, no important aspects of the study have been omitted, and any discrepancies from the study as planned (see preregistration here: https://osf.io/qf49p) have been explained in the supplement.

## Ethical Approval

The UnitedHealth Group Office of Human Research Affairs approved this project and provided the following determination (OHRA Certificate of Action #: 2021-0039) based on the secondary use of de-identified claims data: “The research was determined to qualify as negligible risk and is permissible under exempt category 4 (ii). The research was determined to be exempt as the activities did not identify subjects directly or through identifiers via the claims data in a research database, subjects were not contacted, and investigators will not re-identify subjects.”

## SUPPLEMENTARY MATERIAL

### Data Sources

#### Standardization of Data Entry and Data Structure

Medical and pharmacy claims data are captured, predominantly electronically, from sites of care seeking third-party reimbursement for both Medicare and commercial plans using the industry standard data collection forms HCFA/CMS-1500 for facility claims, UB04/CMS-1450 for professional services and outpatient claims, and NCPDP for pharmacy claims or their electronic equivalents. Structured data from these standardized forms are coded using the International Classification of Diseases, Tenth Revision, Clinical Modification (ICD-10-CM), National Drug Codes (NDC), Current Procedural Terminology (CPT) codes, and Logical Observation Identifiers Names and Codes (LOINC) codes, and Diagnosis Related Groups (DRG). This nomenclature ensures consistency of data collection across geographic regions, health systems, and payers throughout the United States.

#### Methods to Control for Errors in Sampling and Data Collection

Claims that do not adhere to the form or coding standards described above are rejected from reimbursement, minimizing the risk that inappropriately structured data are included in the database.

#### Data Relevance and Accuracy

Data are transferred into the UnitedHealth Group R&D Data Platform, where a dedicated team pursues data management to ensure accurate matching of source data to an individual. This protocol uses unique identifiers to match them to existing identifiers in the UHG R&D Data Platform to determine whether the individual already exists in the platform. A unique identification number is generated for each individual so that data from multiple sources can be linked back to that identification number. Individuals that fail to meet the matching criteria are excluded from the UHG R&D Data Platform to reduce the risk of erroneous linkage of records. Those whose claims do not fulfill basic standardized data structure requirements described previously are also excluded. During this, all member protected data are stored in a separate database that is only accessible by a designated engineering team. In addition to a persistent identifier being generated for each member, a de-identified primary key is also generated. The de-identified primary key is recycled every 6 months, at which time each member is assigned a new de-identified primary key. Data that are made available for research through the UHG R&D Clinical Research Database use the de-identified primary key as the link across data tables. All protected information has been removed, ensuring any research performed is limited to retrospective analysis of de-identified data and accessed in accordance with Health Insurance Portability and Accountability Act regulations.

### Other Drugs for Type 2 Diabetes Treatment

Beyond metformin, there are many other drug classes used to treat type 2 diabetes. The places in therapy for these pharmacologic treatment options are well characterized in randomized controlled trials, with treatment recommendations largely standardized among national and international diabetes care organizations.[15–17] Notably, diabetes guidelines typically recommend using pharmacologic therapies additively, rather than substitutively, and differential recommendations for second-line diabetes drug classes exist for certain subpopulations of people with type 2 diabetes, including for people with or at high risk for atherosclerotic cardiovascular disease (ASCVD), those with heart failure, and those with chronic kidney disease.[15–17] While the variety of available diabetes drug class options may give an appearance of a suite of active comparators for consideration in observational studies, this conclusion ignores the connections to disease severity that certain drug classes may have, as well as the consistency of metformin as a guideline-recommended, first-line therapy in the background of most treatment regimens as other drugs are added over time. A full list of diabetes medications appears in Supplemental Table S6.

### Validation Tools

#### Covariate Balance Diagnostics

Observational studies often attempt to demonstrate that acceptable covariate balance has been achieved between the exposure groups. Whether it comes through matching, inverse propensity weighting, or some other method seeking covariate balance, a demonstration that balance has been achieved is necessary to convince the reader that two groups that are clearly different (as expected by their different prescribed treatments) have been manipulated in such a way that a weighted or reduced sample shows similar covariate distributions on a set of covariates deemed important for minimizing confounding. Figure 1 presents a diagnostic plot showing the pre- and post-adjustment covariate balance achieved in models for the two example outcomes; acceptable covariate balance generally requires the standardized mean differences for all covariates to have absolute values <0.1.

#### Negative Control Experiments

A negative control experiment is one where there exists no causal relationship between the treatment and the outcome.[24] In particular, negative control outcomes, also known as falsification endpoints, preserve the same treatment/control group designations as the primary outcome for each individual in the study, but they are outcomes that cannot reasonably be impacted by the exposure. In our metformin study, outcomes like dry eye syndrome and low back pain are candidate negative control outcomes because there is no known mechanism by which metformin could directly impact these events. Importantly, negative control outcomes should be subject to the same residual confounding as the primary outcomes, which our study assumes to be exclusively related to overall health.

By looking at a wide range of outcomes with no direct, mechanistic connection to the treatment, we seek to expose differences in overall health not accounted for by the treatment or the other observed covariates. For a comparison of metformin and insulin users in a type 2 diabetes cohort, a pattern of nonzero treatment effects for metformin on the negative control outcomes is evidence that we have not adequately controlled for the underlying differences in the overall health status of these two groups.

#### Negative Control Outcome Criteria

The objective in a negative control outcome experiment is to detect significant relationships between the treatment of interest (e.g., metformin) and a mechanistically unrelated outcome (e.g., low back pain); detecting such a relationship raises serious concerns about residual confounding related to the study design. Importantly, these negative control outcomes should be evaluated against these five criteria:

1. There is no mechanistic connection to the treatment under investigation (i.e., no established mechanism of action for this treatment to affect the negative control outcome).
2. Negative control outcomes must be reasonably prevalent; the statistical power associated with the negative control experiment increases as the prevalence of the outcome increases in the population under study. As a rule of thumb, consider giving preference to negative control outcomes at least as prevalent as the primary outcome. A rare negative control outcome will likely be less informative, typically returning a null result even in the presence of significant residual confounding. The noisy results of a large collection of rare negative control outcomes can still be informative, however, even if an individual outcome occurs too rarely to confidently estimate a treatment effect.
3. Negative control outcomes are suspected of being subject to the same residual confounding as the primary outcome under investigation (e.g., does not require a different level of health care access, health insurance benefit design, etc.). This is unverifiable due to the nature of residual confounding, but the point is that whatever may introduce bias in the outcome of interest should be a potential source of similar bias (expected to be the same direction and magnitude) for the negative control outcome.
4. (Optional) There is no causal relationship to disease severity (e.g., not a known indicator of disease severity for the disease indicating this medication).
5. (Optional) The negative control outcome is not an indicator of health-seeking behavior (e.g., some of the most common recorded “diagnoses” in claims data are screenings or exams that could be sex-specific or age-specific). Exceptions are appropriate when the primary outcome is a health-seeking behavior.

Of these five criteria, (1), (2), and (3) are required, and (4) and (5) are desirable in order to further distance the negative outcomes from obvious group differences in disease severity or health care utilization. The entire collection of negative control outcomes will be used to identify meaningful ways in which the comparison groups differ even after adjustments are made for observed covariates. Negative control experiments -- well-accepted and commonly recommended -- remain infrequently conducted components of observational study designs.[23] One explanation for the lack of widespread adoption is that identifying the perfect negative control experiment is often quite challenging. In practice, however, a wide range of negative control experiments do not have to individually be perfect to collectively reveal the residual confounding we seek to expose. For this reason, one might empirically determine a host of acceptable negative control outcomes by reviewing the most frequently observed diagnoses among individuals in the cohort. The focus of this approach is using the collective body of evidence from many possibly imperfect negative experiments rather than relying on any single negative experiment’s ability to survive heavy scrutiny.

#### Negative Control Outcome Selection Algorithm

In a randomized trial, thoughtful selection of negative controls is necessary because that data must be intentionally collected, possibly at significant cost. In an observational study, we must draw candidate negative controls from data that has already been collected as acquiring new data on the selected individuals is highly unlikely. To find outcomes present in our data that meet the criteria for negative controls, we proceed as follows (example results depicted in Table S1):

1. Identify the 500 most common diagnosis codes in 2018 in terms of affected individuals in the primary cohort, ignoring multiple diagnoses for the same condition for the same individual. Repeat for the complementary cohort.
2. Filter the observed diagnoses to only retain diagnoses observed in both cohorts.
3. Rank the diagnosis codes in each cohort and then sum the ranks for a composite rank sum (e.g., low back pain is #13 in prediabetes and #15 in type 2 diabetes for a rank sum of 28).
4. Order the diagnosis codes by rank sums from smallest to largest.
5. Take the top 10/20/50/etc. outcomes that meet the criteria for negative control outcomes. This requires domain expertise to individually consider each outcome for potential mechanistic connections to the treatment, disease severity, and health-seeking behavior.

Additionally, a power analysis can help establish a prevalence minimum. It is likely that these negative control outcomes will span a variety of body systems and will not all be highly correlated, reducing the impact of a single negative control outcome in the group that may have an unrecognized connection to the drug or disease under investigation. Other approaches to identifying negative control outcomes have been developed, including the ATLAS tool created by the Observational Health Data Sciences and Informatics (OHDSI) organization, which also uses a combination of automated discovery and expert review to identify 50-100 negative controls.[23]

**Table S1.** Negative control outcomes were selected through an automated generation of candidate outcomes followed by an expert review. Prevalence of outcomes in both the primary and complementary cohorts is emphasized in this approach, resulting in an ordering of candidates from which experts can identify the first 10/20/50/etc. candidates that satisfy multiple negative control outcome criteria. Here we find three acceptable negative control outcomes in the 10 most common diagnoses. We found 50 suitable negative control outcomes in the ∼100 most prevalent diagnoses in our data set.

| Observed Condition |  | Empirical Prevalence |  |  |  |  | Expert Review |  |  |  |
| --- | --- | --- | --- | --- | --- | --- | --- | --- | --- | --- |
| ICD Code | ICD Code Description | T2D Rank | T2D Total Cases | Prediabetes Rank | Prediabetes Total Cases | Prediabetes and T2D Ranks Summed | well-established mechanistic connection to metformin | impacted by diabetes severity | general health-seeking behavior | negative control candidate |
| i10 | essential (primary) hypertension | 2 | 1419426 | 2 | 146023 | 4 | N | Y | N | N |
| e785 | hyperlipidemia unspecified | 3 | 755895 | 4 | 90418 | 7 | N | Y | N | N |
| z0000 | encounter for general adult medical examination without abnormal findings | 4 | 679139 | 3 | 124587 | 7 | N | N | Y | N |
| z23 | encounter for immunization | 5 | 575125 | 5 | 83550 | 10 | N | N | Y | N |
| e782 | mixed hyperlipidemia | 7 | 479440 | 7 | 55719 | 14 | N | Y | N | N |
| z1231 | encounter for screening mammogram for malignant neoplasm of breast | 8 | 369026 | 6 | 68582 | 14 | N | N | Y | N |
| e039 | hypothyroidism unspecified | 10 | 301838 | 9 | 36737 | 19 | N | N | N | Y |
| r05 | cough | 12 | 271000 | 14 | 30625 | 26 | N | N | N | Y |
| e7800 | pure hypercholesterolemia unspecified | 17 | 248728 | 11 | 34292 | 28 | N | Y | N | N |
| m545 | low back pain | 15 | 262643 | 13 | 31102 | 28 | N | N | N | Y |

#### Complementary Cohorts

Complementary cohorts provide a second tool to stress test the primary results by nullifying or reversing any overall health advantage the treatment group has in the primary cohort. If we suspect the treatment group may be healthier in some unmeasurable way than the comparison group (aside from the possible effect of the treatment), we construct another cohort in which the treatment group is expected to be less healthy than the comparison group (aside from the possible effect of the treatment). The construction of this cohort requires relevant domain expertise in order to satisfy the following criteria:

1. The primary treatment of interest must be reasonably prevalent in the complementary cohort. If there are too few users of the primary treatment in the complementary cohort, there will be limited power to detect an effect.
2. If treatment is concentrated among the healthiest members of the primary cohort (in both measurable and unmeasurable ways), use of the treatment in the complementary cohort should be concentrated among individuals with worse overall health relative to the rest of the cohort, and vice versa. This creates a mirror image of the primary cohort and is critical if the goal is to nullify or reverse the overall health advantage (or disadvantage) suspected in the treated group in the primary cohort.
3. There should be no unnecessary differences in the cohort-identifying disease. This is easiest to satisfy in diseases with a commonly diagnosed “predisease” stage (e.g., prediabetes/type 2 diabetes, albuminuria/chronic kidney disease, osteopenia/ osteoporosis), a framework that best ensures that any residual confounding present in the complementary cohort will be of the same nature as that of the primary cohort (i.e., related to overall health). Introducing a complementary cohort from a completely different disease is still possible, but it may introduce complicated disease differences that must be addressed.

In a study of individuals diagnosed with type 2 diabetes, there is a natural complementary cohort in the population of individuals diagnosed with prediabetes. Critically, the drug metformin is prescribed to individuals in both cohorts -- extensively in type 2 diabetes and more modestly in prediabetes. What makes this primary/complementary cohort specification ideal is where the concentration of metformin users exists in each cohort. In the American Diabetes Association’s (ADA) published “Pharmacologic Approaches to Glycemic Treatment: Standards of Medical Care in Diabetes—2020,” metformin monotherapy is recommended as the first-line treatment for type 2 diabetes along with comprehensive lifestyle modifications.[15] If comorbidities like atherosclerotic cardiovascular disease, heart failure, or chronic kidney disease are present, other drugs may augment or replace metformin. If metformin and/or other drugs cannot effectively control blood glucose, an individual may ultimately be prescribed insulin. Thus, individuals with a metformin claims history, but no history of insulin use, are earlier in the spectrum of type 2 diabetes severity than those who have progressed to using insulin.

In prediabetes, the ADA Standards of Medical Care recommend considering treatment with metformin for individuals at risk for developing type 2 diabetes, particularly for high-risk individuals, including those with a history of gestational diabetes, BMI >= 35 kg/m^2^, or age less than 60 years old.[14] Whereas the metformin users were the least severe cases in the type 2 diabetes cohort, they hold the opposite position in the prediabetes cohort. As depicted in Figure S1, prediabetes thus presents an ideal complementary cohort by eliminating or potentially reversing any metformin exposure group advantage that could be attributable to overall patient health in the primary cohort.

**Figure S1.**
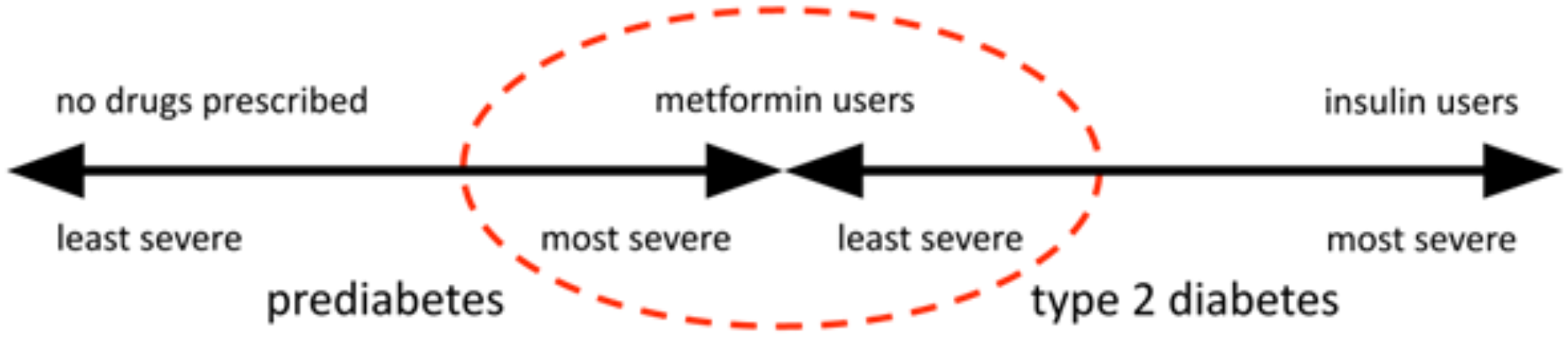
Based on guideline-driven treatment recommendations, metformin users diagnosed with prediabetes are assumed to be on the opposite end of their respective diabetes severity spectrum compared to metformin users diagnosed with type 2 diabetes. The prediabetes cohort thus reverses the overall health advantage enjoyed by the metformin users in the type 2 diabetes cohort, which makes it an ideal candidate for a complementary cohort aiming to expose residual confounding related to overall health.

The role of this complementary cohort is to validate whether the covariate selections and method choices in the primary cohort analysis are indeed effective at addressing confounding related to disease severity and overall health. If they are, we should expect to reproduce the primary cohort finding in the complementary cohort. Conflicting findings suggest that the result in the primary cohort may be a result of residual confounding and not a true treatment effect. Other explanations exist for conflicting findings, specifically a heterogeneous treatment effect, a possibility that goes unexplored when the entire focus is on the primary cohort.

#### Negative Controls in Complementary Cohorts

Negative control outcome results may reveal residual confounding in the primary cohort (e.g., through a pattern of protective associations between the treatment and the negative control outcomes), which further motivates the use of a complementary cohort to test the primary result. These same negative control outcomes must also test the selection of the complementary cohort, ensuring it exhibits the most desired quality of a complementary cohort: no residual confounding-induced advantage in negative control outcome experiments. The strength of a complementary cohort is defined by how much the negative control outcome associations are nullified or reversed in comparison with the primary cohort results. Larger reversals indicate that the complementary cohort provides a more robust validation of the initial results. If the pattern of bias (i.e., nonzero negative control outcome effect sizes) is similar in the two groups, then the second analysis has little to add aside from validating the result in another population.

When adequately powered, null results across all negative control outcome experiments in both the primary and complementary cohorts are a good indication that residual confounding may be fairly minimal in the identified cohorts and the results are likely trustworthy. When the primary and complementary cohorts yield conflicting results across a host of negative control outcomes, we attribute that difference to a difference in residual confounding in the two cohorts -- this is exactly what we hope to uncover if it exists. In Figure 3, we capture these residual confounding differences in an easily digestible diagnostic plot; it is this diagnostic plot that provides the necessary backdrop to interpret a study’s primary result from a more informed position.

#### Covariate Balance in Negative Control Outcome Experiments

Covariate balance was demonstrated for the primary outcomes in the primary and complementary cohorts in the main text. In Figure S2 we show that acceptable covariate balance was achieved using IPW for 50 negative control outcome experiments conducted in both cohorts.

**Figure S2.**
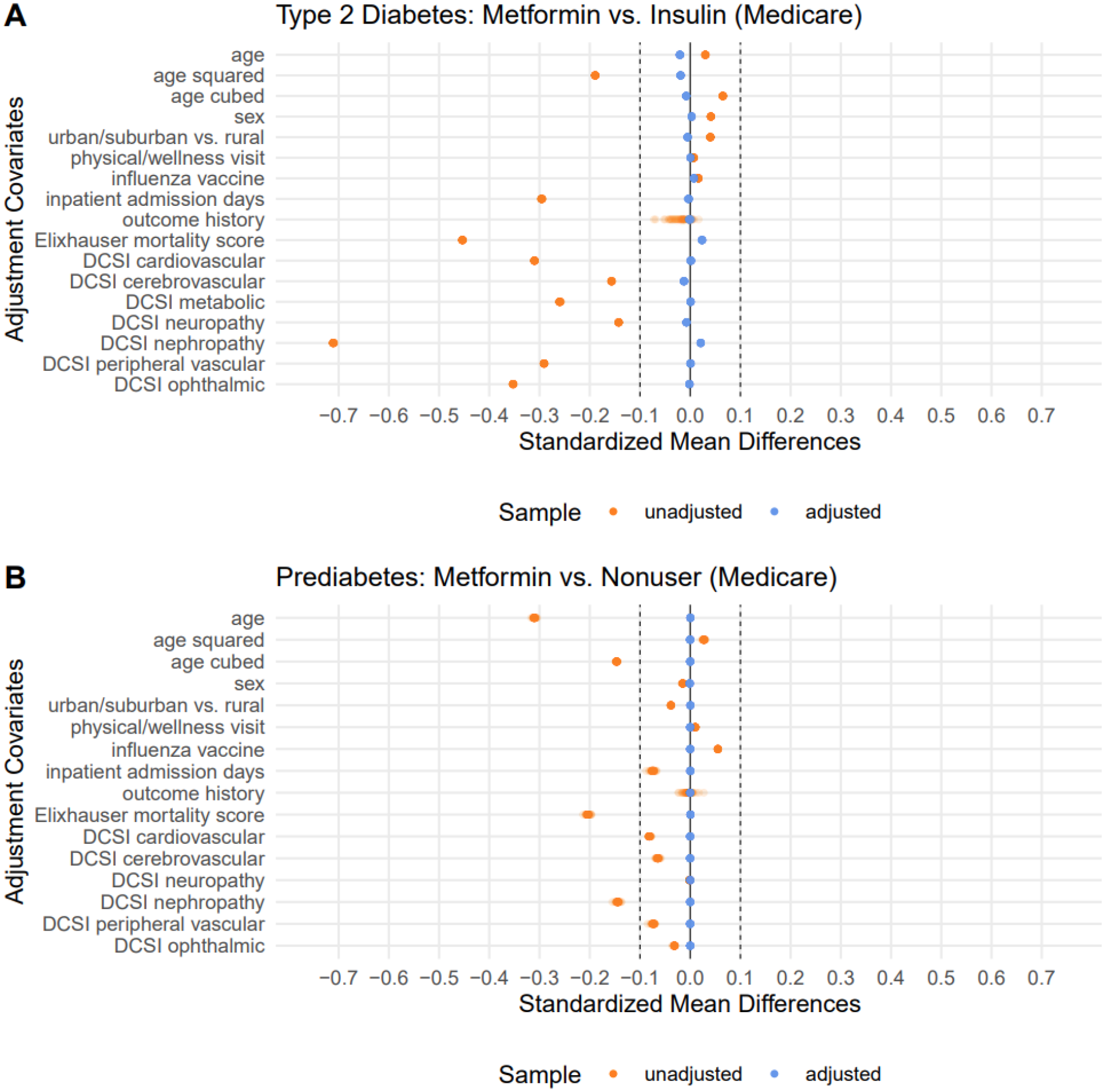
Here we show 50 overlaid balance plots for the Medicare Advantage type 2 diabetes cohort (panel A) and the prediabetes cohort (panel B) where the only covariate changing from one negative control outcome experiment to the next is the history of each respective outcome. The fact that adjusted balance changes negligibly with the exception of outcome history suggests that the sample under analysis (after propensity trimming) is largely the same from one experiment to the next. The noticeable leftward shift in unadjusted balance in panel A reflects the healthier nature of the metformin group in the type 2 diabetes cohort (fewer inpatient days, lower Elixhauser in-hospital mortality score, lower prevalence of diabetes complications in every DCSI dimension despite being slightly older as a group). In panel B, we see better unadjusted balance that appears to meaningfully reduce the metformin advantage. Across both cohorts, the IPW approach achieved satisfactory balance after adjustment for every observed covariate of interest.

#### Final Results Plot Interpretation

Figure 4 in the main text (as well as Figure S7, Figure S11, and Figure S15 in the supplement) depicts the primary and complementary cohorts results for a primary outcome against a backdrop of negative control outcome experiment results. The discussion surrounding these figures presents an interpretation for each figure, but not all possible scenarios were observed in the real data example. In Figure S3 we explore a more comprehensive set of possible findings that may appear in a main results figure, and we provide a recommended interpretation for each set of results. The interpretation of each scenario centers on examining each cohort’s primary result in the context of its negative control outcome distribution; we then check for agreement between the cohorts.

**Figure S3.**
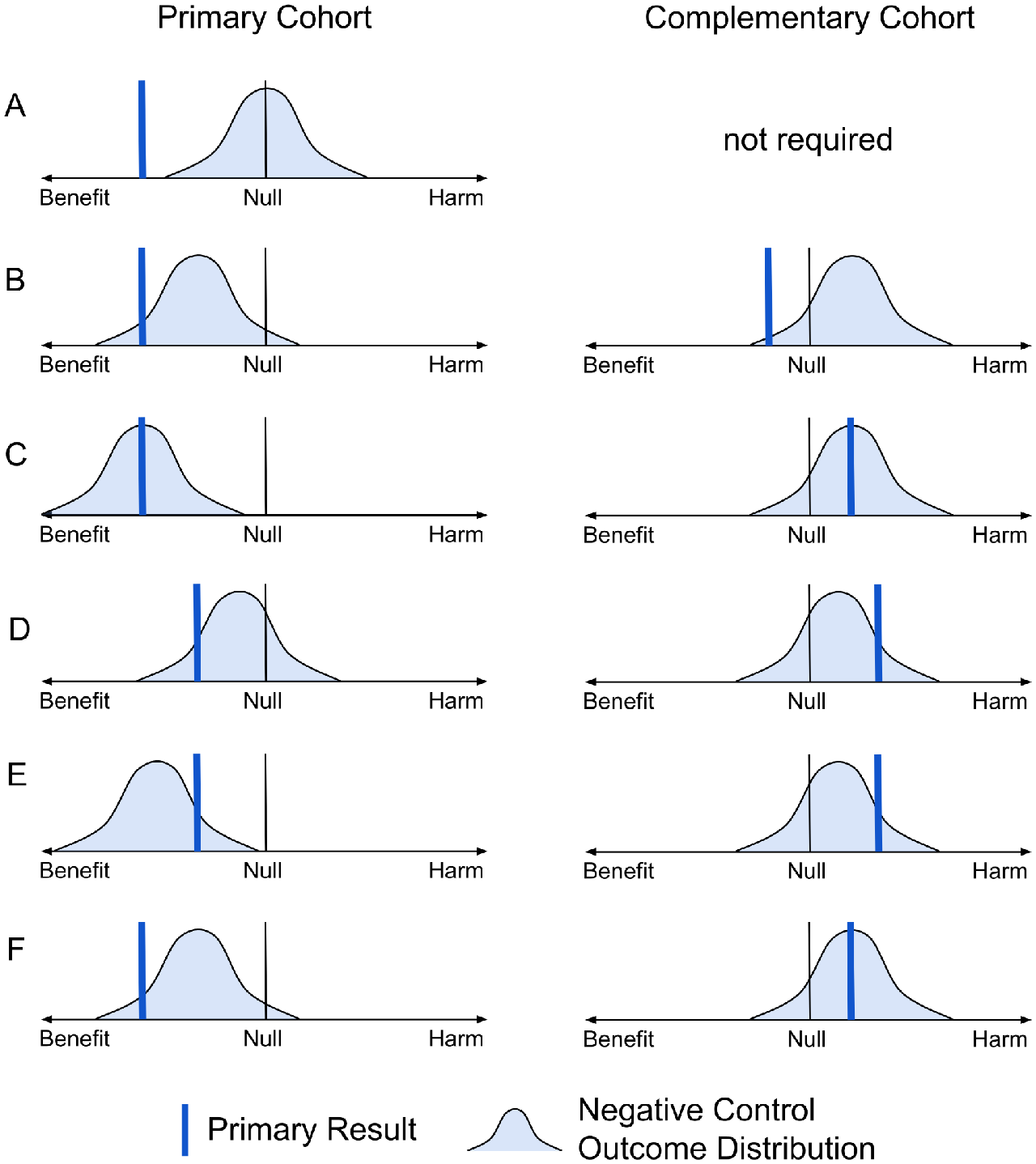
Possible Primary Outcome Results: In each of these scenarios, a primary cohort is depicted with a blue bar identifying the result for the primary outcome, and a light blue distribution of negative control outcome results appears behind it. The complementary cohort results are depicted in the same manner. The discussion in the supplement text examines each row A-F for the level of evidence it provides to support a claim of a beneficial treatment effect.

In Figure S3 row A, we see a strong primary result and no pattern of bias in the negative control outcome experiments. With no evidence of bias, there is no requirement to have a complementary cohort, and we have reason to trust the primary result. We would similarly trust a result indicating harm the farther right it is from the null. Our confidence in either result would decrease as it moves toward the null.

In Figure S3 row B, we observe a strong primary result, but there is a clear pattern of bias in the negative control outcome experiments, which weakens the evidence supplied by the primary result. In the complementary cohort (validated by observed bias reversal), we see a more modest primary result, but it is quite strong compared to the negative control outcome experiments. Taken together, all the evidence from both cohorts points to a beneficial treatment effect. In this case, the complementary cohort analysis strengthened our confidence in the primary cohort result.

In Figure S3 row C, a strong primary result is nullified by the negative control outcome experiments. An unfavorable result in the complementary cohort is also nullified by negative control outcome experiments. Taken together, there is no strong evidence of any effect.

In Figure S3 row D, we see a modest primary result in the primary cohort, and this result exceeds a large majority of the negative control outcome experiments. In the complementary cohort, everything is reversed such that the primary result is now harmful and exceeds a similarly large majority of the negative control outcome experiments. Taken together, these conflicting findings present no strong evidence of any effect. In this case, the complementary cohort analysis erased our confidence in the primary cohort result.

In Figure S3 row E, a modest primary result in the primary cohort actually appears harmful compared to the distribution of negative control outcome experiments. In the complementary cohort, the result indicating harm is worse than a large majority of the negative control outcome distribution. Taken together, these results suggest there may be a harmful effect. In this case, the complementary cohort analysis strengthened our confidence in the primary cohort result.

In Figure S3 row F, a strong primary result in the primary cohort exceeds a large majority of the negative control outcome results, indicating a potentially beneficial treatment effect. In the complementary cohort, a result indicating harm is squarely in the middle of the negative control outcome results, effectively indicating a null effect. Taken together, these results are inconclusive. There’s some evidence supporting benefit and other evidence suggesting no effect. In this case, the complementary cohort analysis lessened our confidence in the primary cohort result.

### Supplementary Analyses

The main text focused on a comparison group of insulin users, but nonusers are also a frequent comparison group in studies of metformin and other drugs. Nonusers are a difficult group to conceptualize when virtually every stage of treatment for a condition involves prescription medication (as seen in the type 2 diabetes treatment recommendations). The “nonuser” population can also be hard to describe when the drug under investigation is available inexpensively without using insurance (e.g., metformin). Since metformin is so widely prescribed in the type 2 diabetes population, there is a reasonable chance that some “nonusers” are taking metformin; they are simply purchasing it outside the visibility of their insurance plans, making them only appear as nonusers in our study despite obtaining the medication through alternate means such as cash pay. This has the effect of biasing any effect estimate toward the null and was the primary reason we selected insulin users as the comparison group for the main analysis.

In Tables S2-S4 and Figures S4-S115 we show three supplemental analyses not presented in the main text. Each of the three analyses is presented in one results table and four figures. Table S2, Figure S4, Figure S5, Figure S6, and Figure S7 present a metformin analysis in the Medicare Advantage population with a comparison group of nonusers (different comparison group from the main text). Table S3, Figure S8, Figure S9, Figure S10, and Figure S11 present a metformin analysis in the commercially insured population with a comparison group of insulin users (different population from the main text). Table S4, Figure S12, Figure S13, Figure S14, and Figure S15 present a metformin analysis in the commercially insured population with a comparison group of nonusers (different population and comparison group from the main text). Each four-figure group shows two Love plot figures (one for the example outcomes and one for the 50 negative control outcome experiments) depicting acceptable covariate balance, a residual confounding plot, and finally the primary outcome results plotted on a distribution of negative control outcome effect estimates in both the primary and complementary cohorts. We see bias in the same direction emerge in the distributions of negative control outcome effect estimates in every combination of population and comparison group definition. In summary, none of the populations and comparison groups we explored appear immune to a concerning amount of residual confounding.

#### Medicare Advantage Beneficiaries (metformin users vs. nonusers)

Examining the nonuser comparison group in the Medicare Advantage population produced favorable results supporting a metformin benefit (see Table S2). The large Medicare Advantage population makes these conservative results highly confident. The pre-adjustment covariate balance in Figure S4 and Figure S5 indicates a slightly healthier metformin user population, and Figure S6 confirms once again through the negative control outcome experiments that a strong bias exists favoring metformin in type 2 diabetes while also showing a weaker bias against metformin in the prediabetes cohort. Interestingly, Figure S7 shows that the example outcome effect estimates in type 2 diabetes are at best as strong as those seen for an average negative control outcome experiment, but they are far worse than an average negative control outcome experiment in the prediabetes cohort. Thus, while Table S2 may indicate a favorable treatment effect estimate, no such conclusion can be supported by the total evidence supplied by the complementary cohort design.

**Table S2.** Treatment effect estimates for metformin: this study was conducted in a Medicare Advantage type 2 diabetes population comparing metformin users to a control group of nonusers. The outcomes represent >0 inpatient admission days in 2019 and a total medical spend (insurance payouts to health care providers) exceeding the 90th percentile of all type 2 diabetes patient expenditures (>$25,793). Metformin appears associated with fewer inpatient admission days and lower health care costs, even after adjustment for a range of relevant covariates.

| outcome | model | log OR (base 2) | 95% CI | p | E-value |
| --- | --- | --- | --- | --- | --- |
| inpatient days | unadjusted | -0.39 | (-0.41, -0.36) | $<10^{-216}$ | 1.94 |
| inpatient days | IPW logistic | -0.09 | (-0.12, -0.06) | $<10^{-10}$ | 1.33 |
| medical spend | unadjusted | -0.55 | (-0.58, -0.53) | $<10^{-323}$ | 2.30 |
| medical spend | IPW logistic | -0.17 | (-0.20, -0.14) | $<10^{-23}$ | 1.50 |

**Figure S4.**
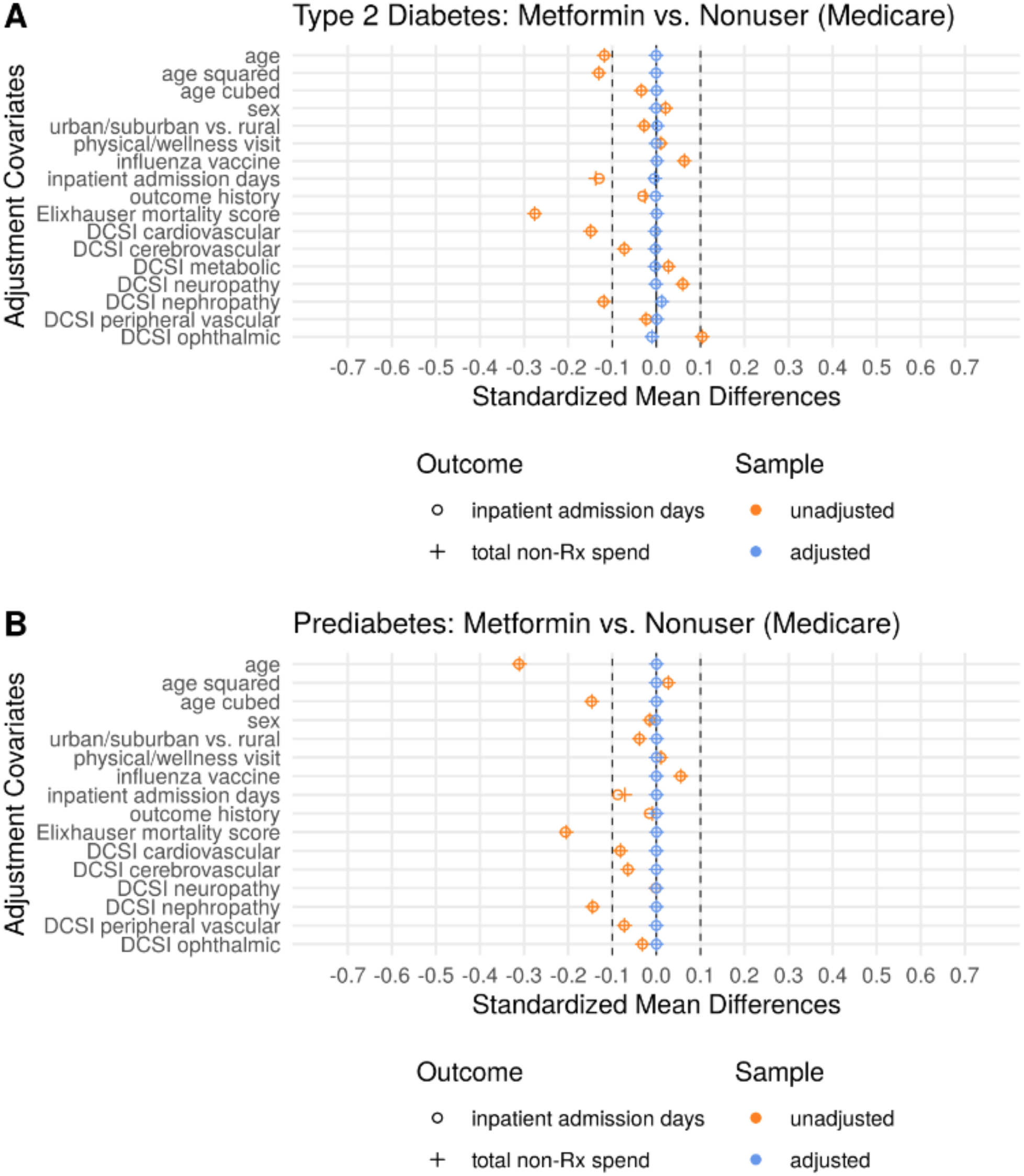
Covariate Balance in Example Outcomes: Medicare Advantage Beneficiaries (metformin users vs. nonusers). This is a different comparison group compared to Figure 1, and in both the prediabetes and type 2 diabetes cohorts, there appears to be a slight health advantage among the metformin users (pre-adjustment standardized mean differences <0). In all cases, the post-adjustment balance is excellent.

**Figure S5.**
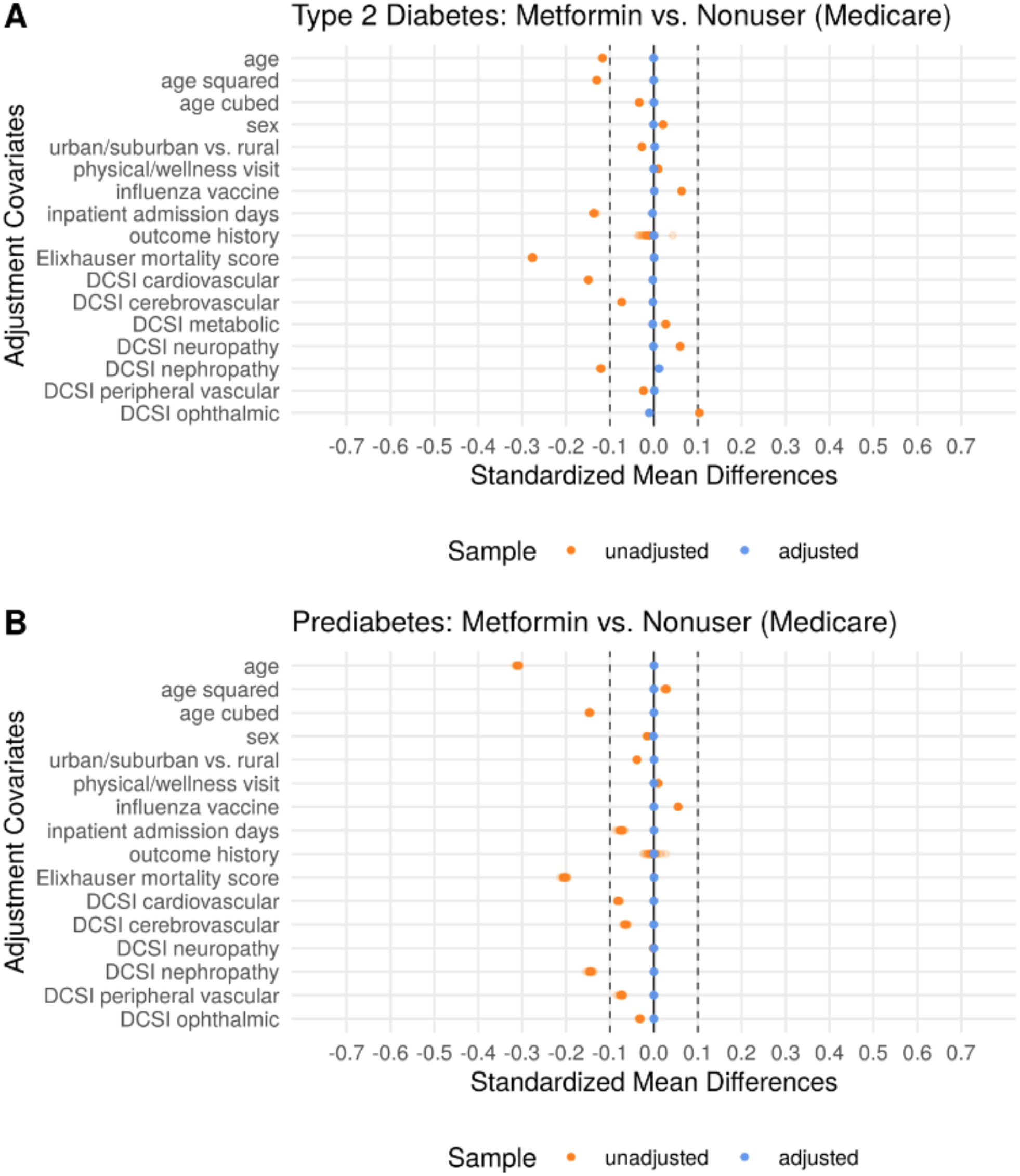
Covariate Balance in Negative Control Outcome Experiments: Medicare Advantage Beneficiaries (metformin users vs. nonusers). This is a different comparison group compared to Figure S2, and in both the prediabetes and type 2 diabetes cohorts, there appears to be a slight health advantage among the metformin users (pre-adjustment standardized mean differences <0). In all cases, the post-adjustment balance is excellent.

**Figure S6.**
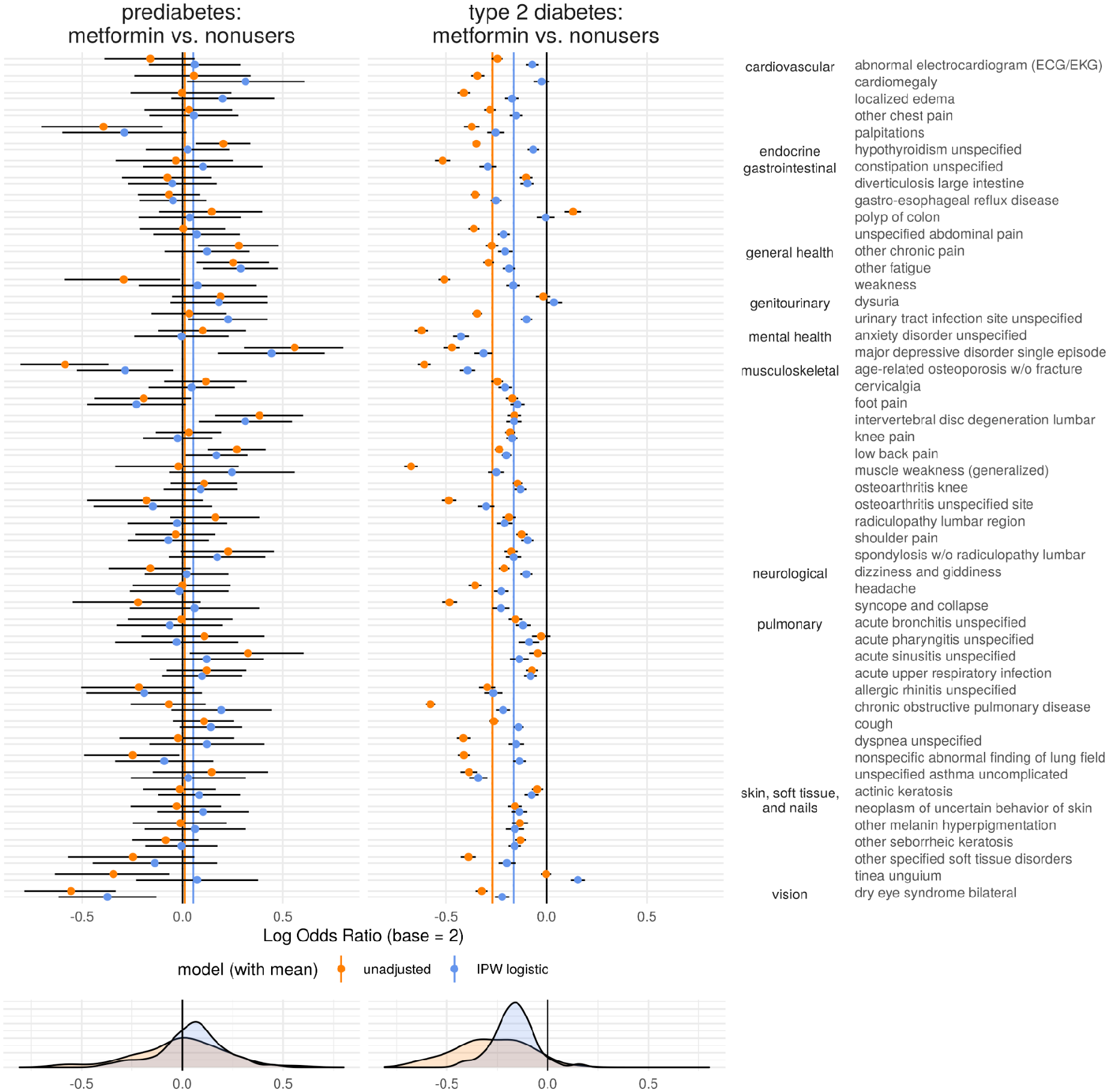
Residual Confounding Plot: Medicare Advantage Beneficiaries (metformin users vs. nonusers of any diabetes drug). This is a different comparison group compared to Figure 3 in the main text. The residual confounding again appears consistent with the primary analysis, strongly favoring metformin users in the type 2 diabetes cohort while maintaining a smaller bias against metformin in the prediabetes cohort.

**Figure S7.**
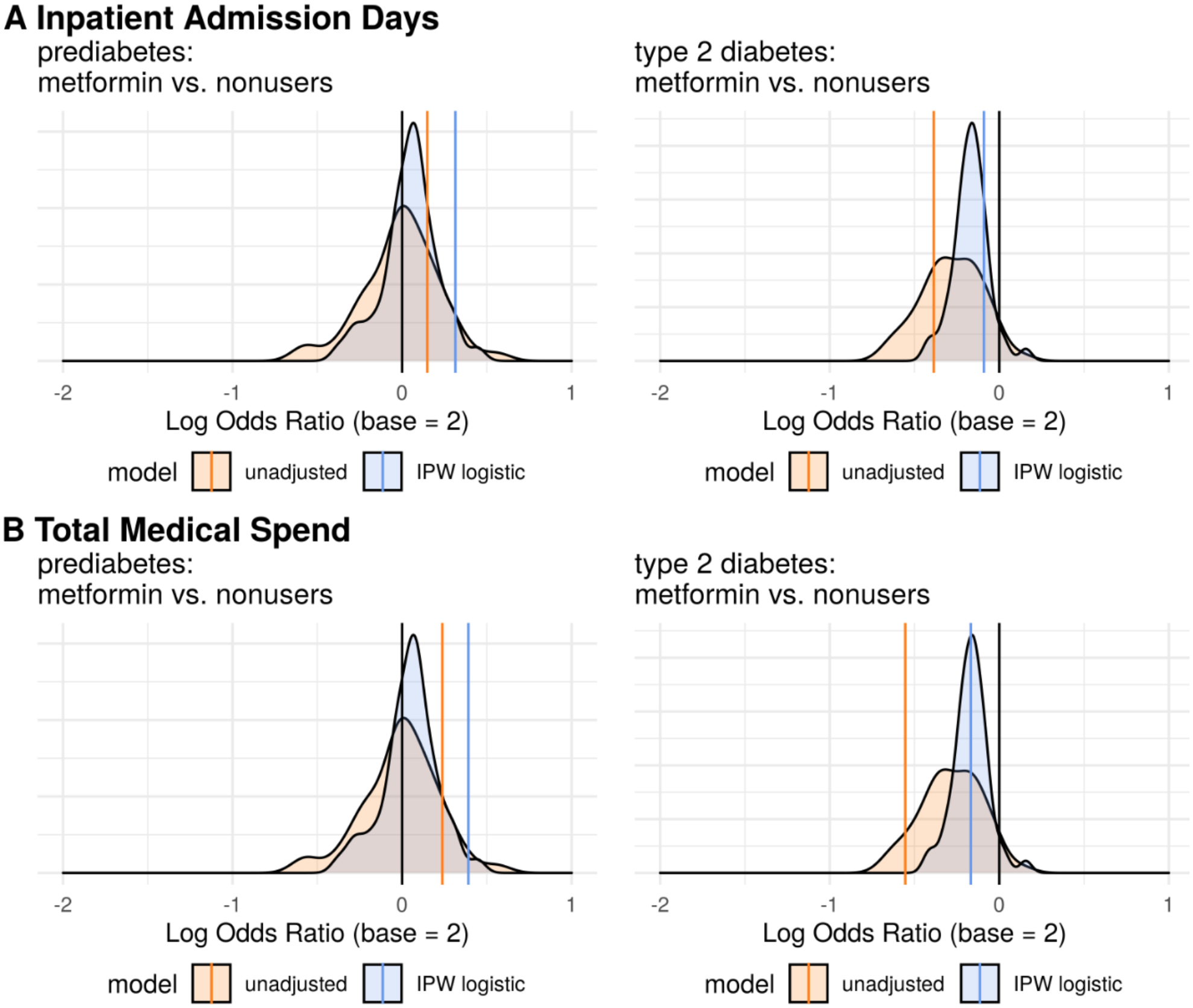
Primary Outcome Results: Medicare Advantage Beneficiaries (metformin users vs. nonusers of any diabetes drug). This is a different comparison group compared to Figure 4 in the main text. We see adjusted treatment effect estimates that are at best on par with an average effect size for a negative control outcome in type 2 diabetes and considerably worse in prediabetes; together, these observations should elicit doubt about any claims of a real effect in the primary analysis.

#### Commercial Insurance Beneficiaries (metformin users vs. insulin users)

In an alternate population of commercially insured beneficiaries, we see strong results in Table S3 favoring metformin usage that are quite confident despite the much smaller population under study. Figure S8 and Figure S9 show excellent covariate balance, but we continue to see in Figure S10 negative control outcome effect estimates biased in favor of metformin in the type 2 diabetes population and biased against metformin in the prediabetes population. Interestingly, while the adjusted effect estimates for the example outcomes indicate a potential treatment effect that appears relatively strong compared to the negative control outcome effect estimates in Figure S11, one effect goes to 0 while the other substantially reverses in the prediabetes population. Together, these results suggest that the findings in Table S3 are likely products of significant residual confounding.

**Table S3.** Treatment effect estimates for metformin: this study was conducted in a commercially insured type 2 diabetes population comparing metformin users to a control group of insulin users. The outcomes represent >0 inpatient admission days in 2019 and a total medical spend (insurance payouts to health care providers) exceeding the 90th percentile of all type 2 diabetes patient expenditures (>$21,433). Metformin appears strongly associated with fewer inpatient admission days and lower health care costs, even after adjustment for a range of relevant covariates.

| outcome | model | log OR (base 2) | 95% CI | p | E-value |
| --- | --- | --- | --- | --- | --- |
| inpatient days | unadjusted | -1.49 | (-1.62, -1.36) | $<10^{-106}$ | 5.07 |
| inpatient days | IPW logistic | -0.71 | (-0.89, -0.52) | $<10^{-13}$ | 2.65 |
| medical spend | unadjusted | -1.51 | (-1.63, -1.40) | $<10^{-134}$ | 5.15 |
| medical spend | IPW logistic | -0.65 | (-0.82, -0.48) | $<10^{-13}$ | 2.51 |

**Figure S8.**
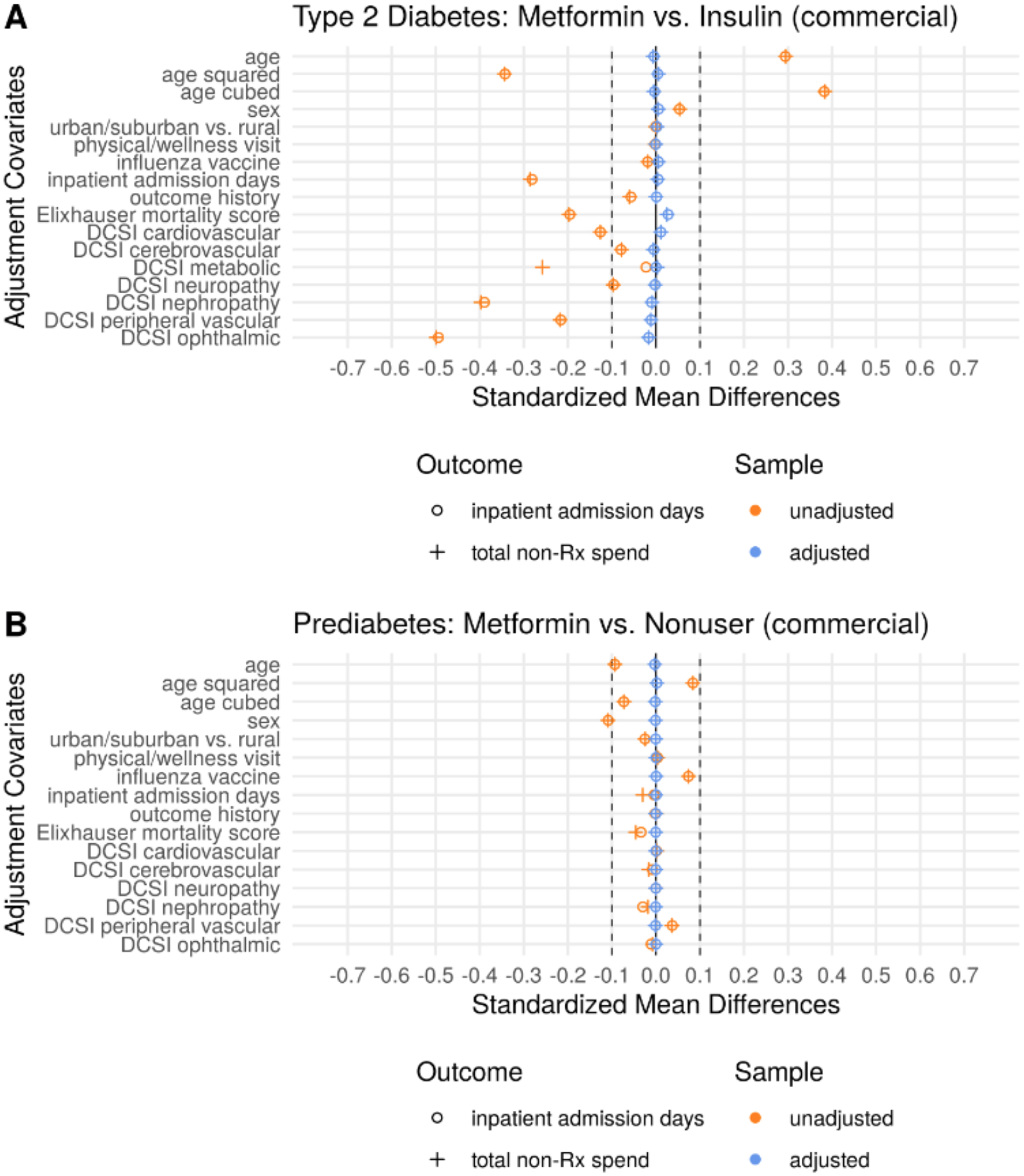
Covariate Balance in Example Outcomes: Commercially Insured Beneficiaries (metformin users vs. insulin). This is a different population compared to Figure 1. In the type 2 diabetes cohort, the metformin users have a noticeable health advantage that essentially disappears in the prediabetes population. In both the prediabetes and type 2 diabetes cohorts, the post-adjustment balance is excellent.

**Figure S9.**
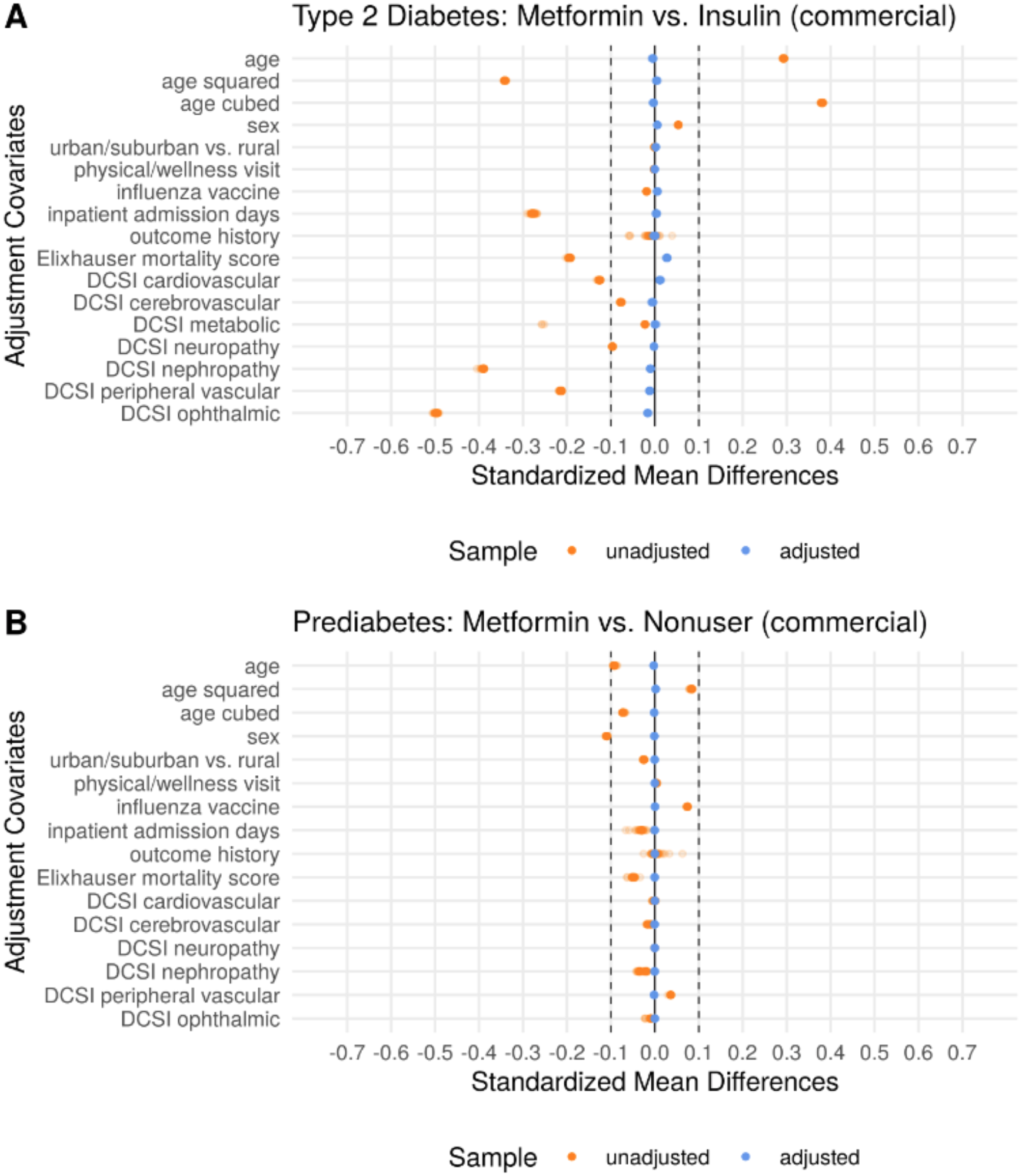
Covariate Balance in Negative Control Outcome Experiments: Commercial Insurance Beneficiaries (metformin users vs. insulin users). This is a different population compared to Figure S2. Here we show 50 overlaid balance plots for the type 2 diabetes cohort (panel A) and the prediabetes cohort (panel B) where the only covariate changing from one negative control outcome experiment to the next is the history of each respective outcome. Excellent post-adjustment covariate balance is achieved for all negative control outcome experiments.

**Figure S10.**
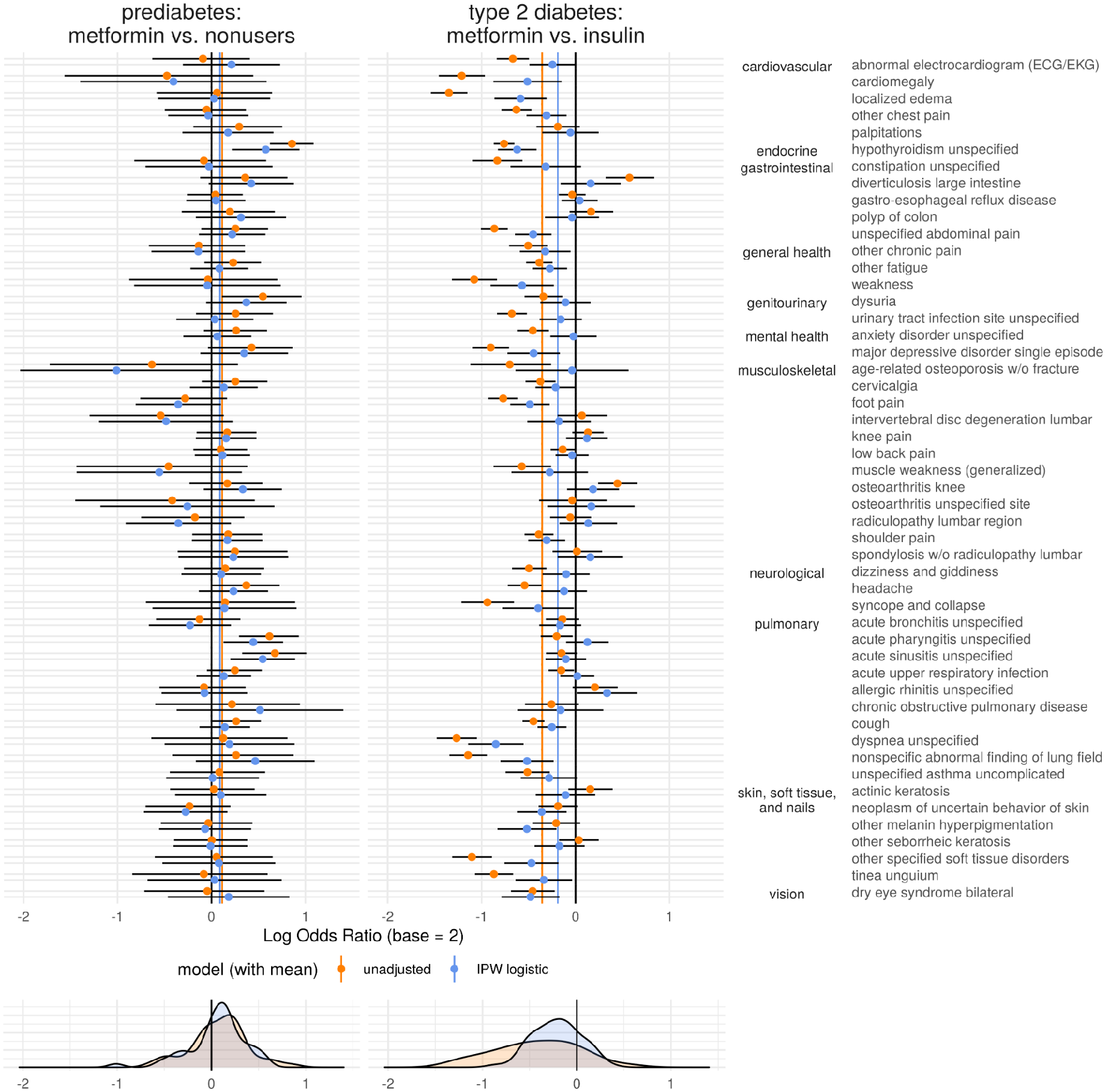
Residual Confounding Plot: Commercially Insured Beneficiaries (metformin users vs. insulin users). This is a different population compared to Figure 3 in the main text. The residual confounding again appears consistent with the primary analysis, strongly favoring metformin users in the type 2 diabetes cohort while appearing to work against metformin users in the prediabetes cohort.

**Figure S11.**
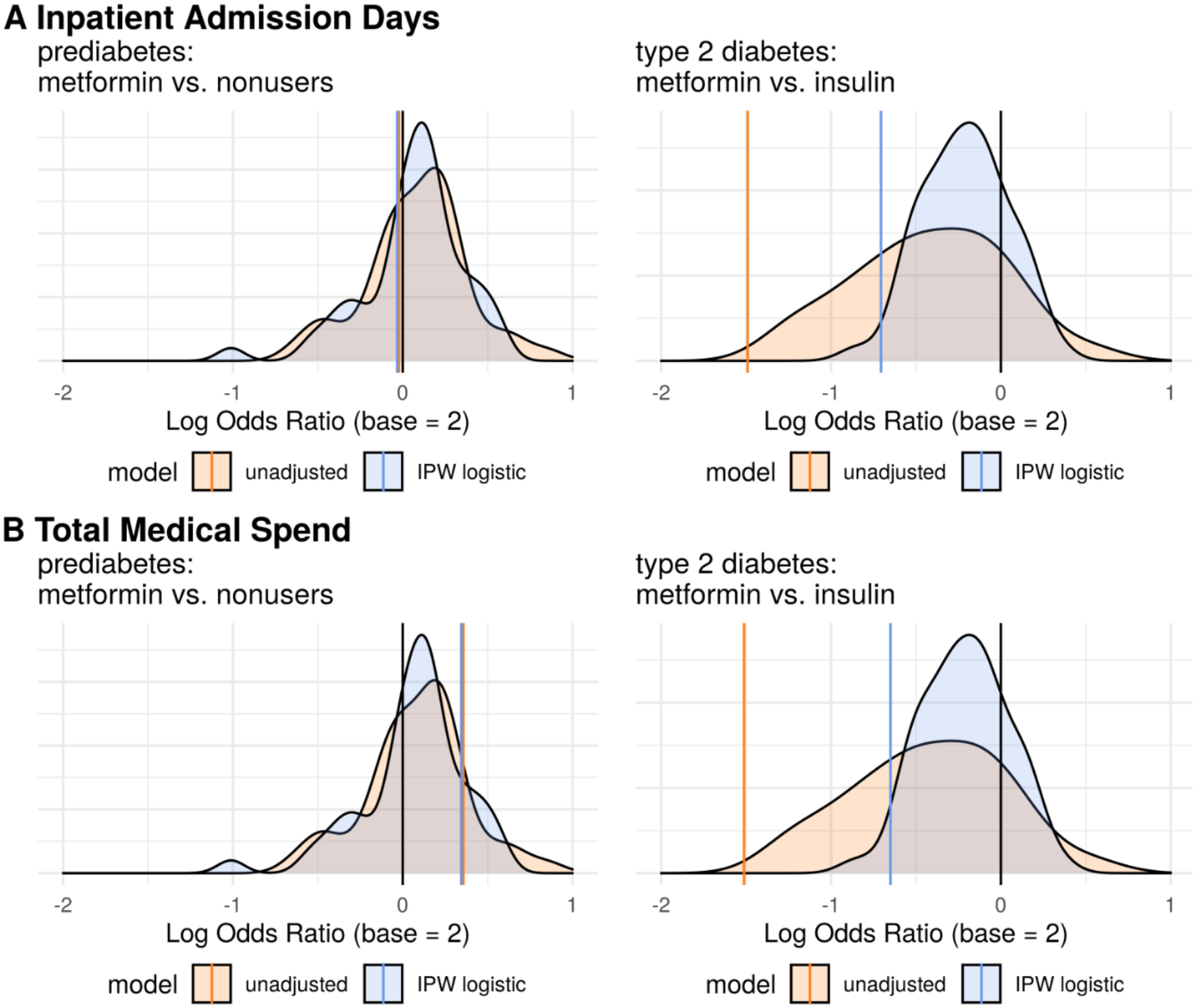
Primary Outcome Results: Commercially Insured Beneficiaries (metformin users vs. insulin users). This is a different population compared to Figure 4 in the main text. We see adjusted treatment effect estimates that exceed a large majority of the estimated effect sizes from the negative control outcome experiments in type 2 diabetes. In prediabetes, however, these effect estimates become null in one case and substantially reverse in the other, which together should elicit some doubt about any claims in the primary analysis.

#### Commercial Insurance Beneficiaries (metformin users vs. nonusers)

In an alternate comparison group of nonusers of any diabetes drugs among the commercially insured, we see more conservative results favoring metformin usage in Table S4. While the nonusers may be hard to completely explain, we can be relatively confident they are not insulin users (and thus more advanced type 2 diabetes cases) due to the generally high list price of insulins. Figure S12 and Figure S13 show excellent covariate balance, but we continue to see in Figure S14 negative control outcome effect estimates biased in favor of metformin in the type 2 diabetes population and biased against metformin in the prediabetes population. Interestingly, while the adjusted effect estimates for the example outcomes indicate a potential treatment effect with varying levels of confidence, neither type 2 diabetes effect estimate is stronger than even half of the negative control outcome effect estimates in Figure S15. This observation strongly challenges the results in Table S4 as nothing more than products of significant residual confounding.

**Table S4.** Treatment effect estimates for metformin: this study was conducted in a commercially insured type 2 diabetes population comparing metformin users to a control group of nonusers. The outcomes represent >0 inpatient admission days in 2019 and a total medical spend (insurance payouts to health care providers) exceeding the 90th percentile of all type 2 diabetes patient expenditures (>$21,433). Metformin appears strongly associated with fewer inpatient admission days and lower health care costs, even after adjustment for a range of relevant covariates.

| outcome | model | log OR (base 2) | 95% CI | p | E-value |
| --- | --- | --- | --- | --- | --- |
| inpatient days | unadjusted | -0.39 | (-0.49, -0.29) | $<10^{-13}$ | 1.95 |
| inpatient days | IPW logistic | -0.15 | (-0.26, -0.04) | $<0.01$ | 1.45 |
| medical spend | unadjusted | -0.26 | (-0.35, -0.17) | $<10^{-7}$ | 1.69 |
| medical spend | IPW logistic | -0.09 | (-0.19, 0.01) | 0.08 | 1.33 |

**Figure S12.**
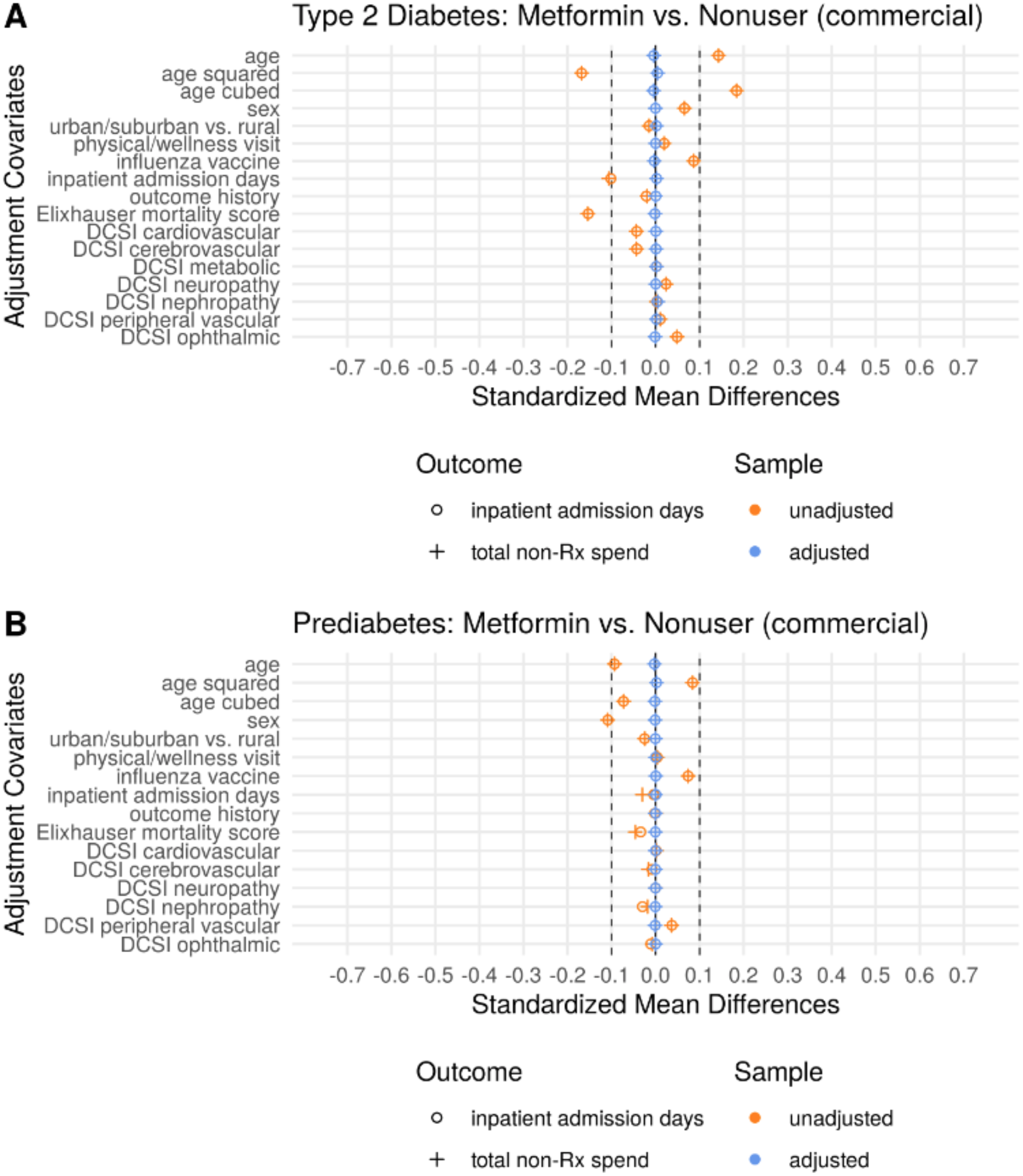
Covariate Balance in Example Outcomes: Commercial Insurance Beneficiaries (metformin users vs. nonusers). This is the same analysis as Figure 1 in the main text, but it considers a different population and comparison group. Compared to Figure 1, the unadjusted balance in the type 2 diabetes population indicates metformin users are much more comparable to nonusers than insulin users.

**Figure S13.**
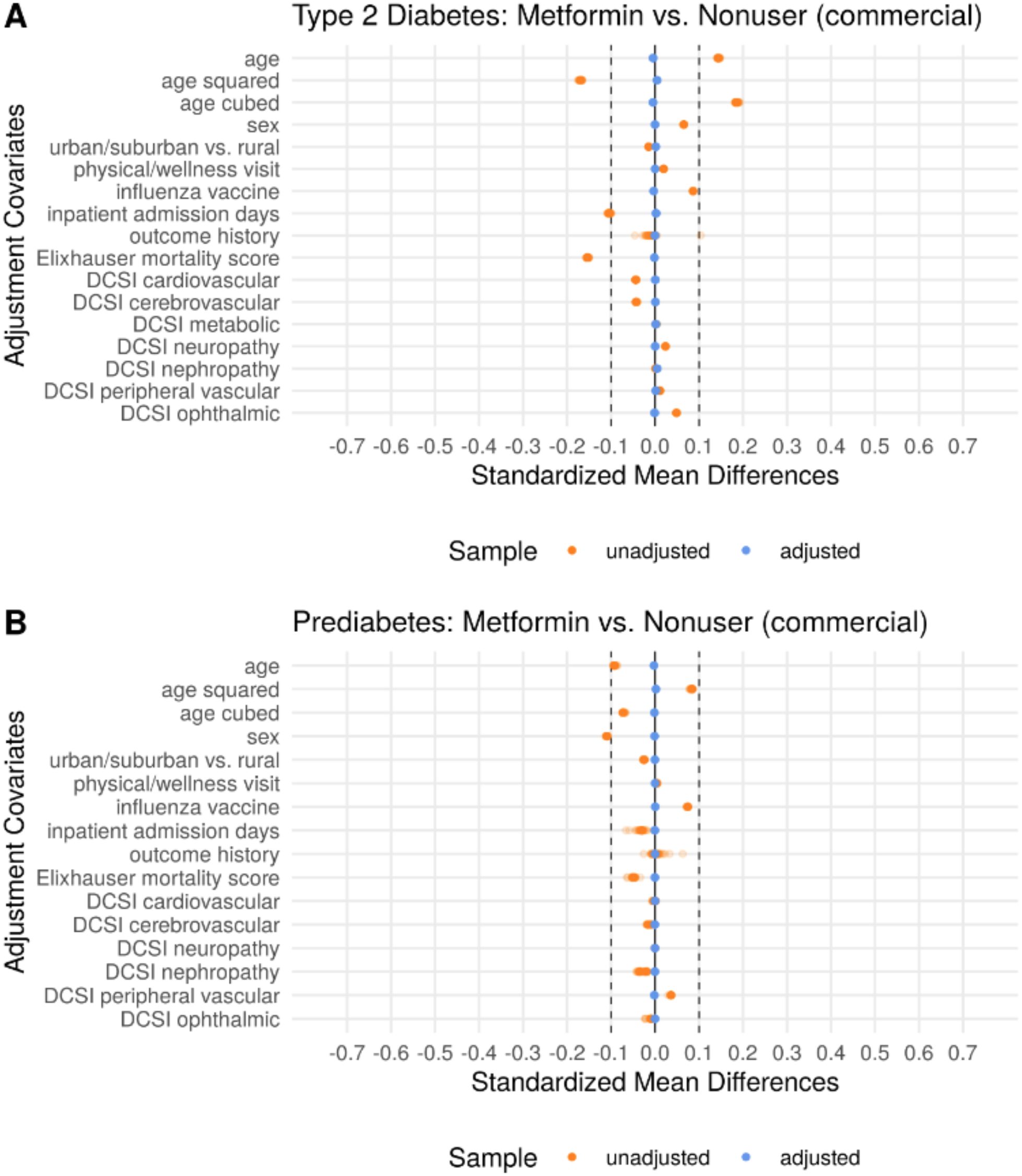
Covariate Balance in Negative Control Outcome Experiments: Commercial Insurance Beneficiaries (metformin users vs. nonusers). This is a different population and comparison group compared to Figure S2. The type 2 diabetes metformin users and nonusers are considerably better balanced pre-adjustment here compared to Figure S2, but that is not enough to eliminate the bias we see in Figure S14.

**Figure S14.**
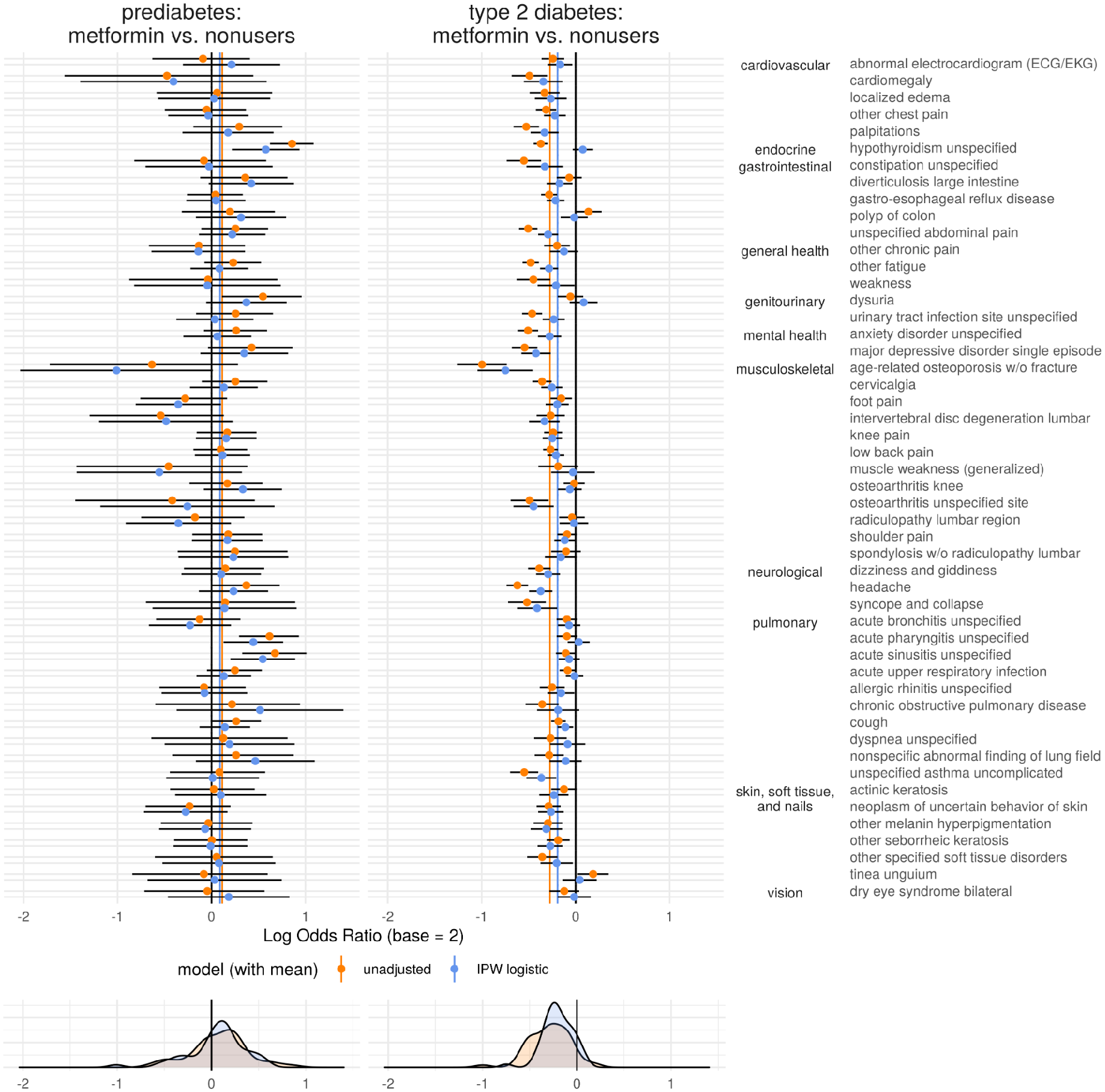
Residual Confounding Plot: Commercial Insurance Beneficiaries (metformin users vs. nonusers of any diabetes drug). This is the different population and comparison group compared to Figure 3 in the main text. The effect sizes are more conservative with this comparison group (though still biased to favor metformin in type 2 diabetes and oppose metformin in prediabetes), possibly due to the number of metformin users mixed into the nonuser population.

**Figure S15.**
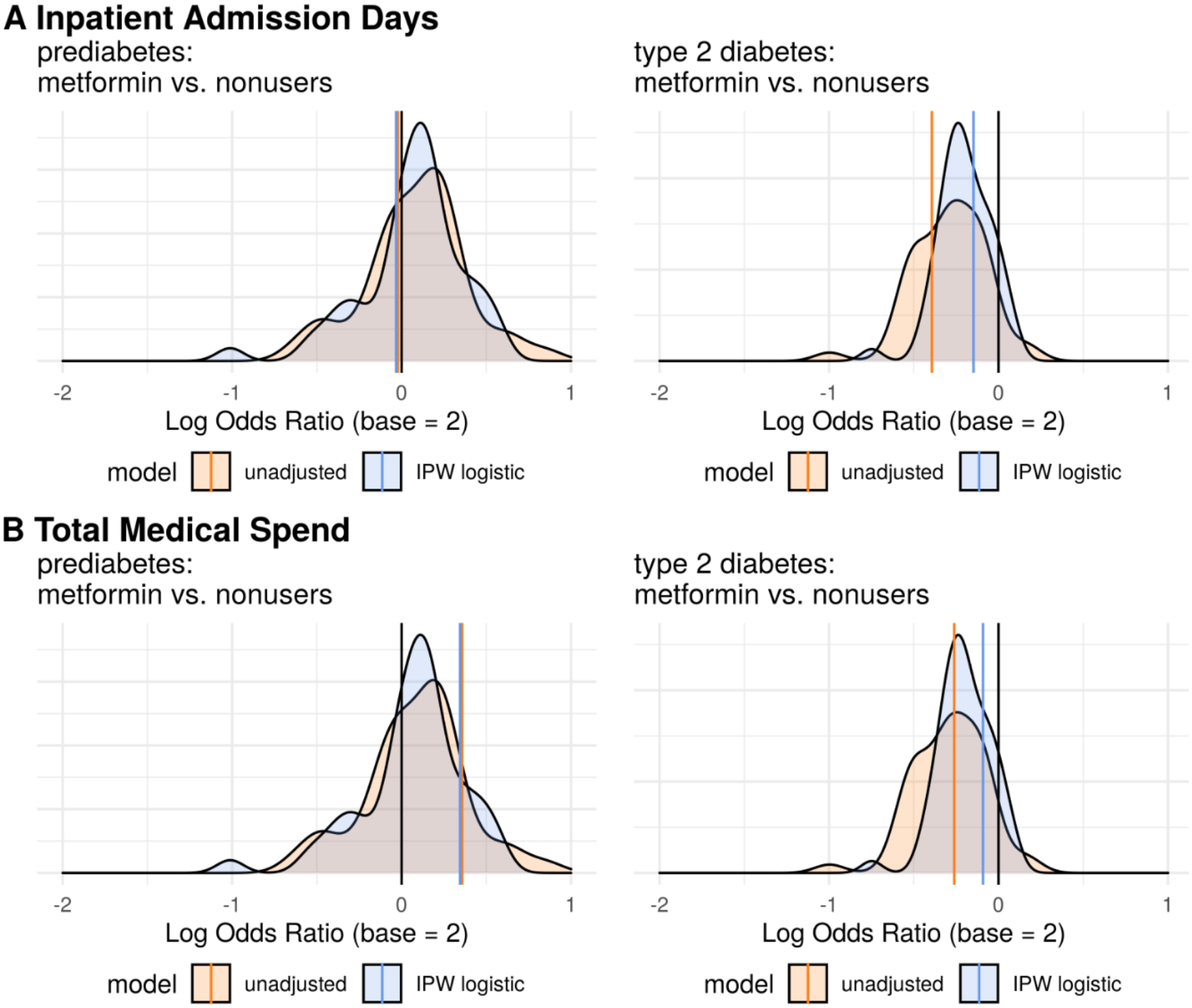
Primary Outcome Results: Commercial Insurance Beneficiaries (metformin users vs. nonusers of any diabetes drug). This is a different population (commercially insured) compared to Figure 4 in the main text, and it considers a different type 2 diabetes comparison group: nonusers. In this population and comparison group setting, the adjusted effect estimates in the type 2 diabetes setting are not even as favorable as what we observe for an average negative control outcome, which is an immediate indicator that the observed association may be spurious.

#### Deviations from Preregistration

The preregistered analysis plan for this study can be found at https://osf.io/qf49p.

Deviations from this plan are listed and explained below:

1. Covariates - The preregistration states that “health-seeking behavior will be indicated by the presence of at least one immunization (typically a flu shot).” We instead used *only* flu shots because of the widespread eligibility and anticipated annual frequency of flu shots not common to all vaccinations.
2. Outcomes - We did not specify any primary outcomes for the example analysis in the preregistration. We realized after conducting all the negative control experiments that we were missing the opportunity to illustrate interpreting a real result. We only ever tried two example outcomes, and both are reported in the four analyses spanning the main text and supplement.

As the stated criteria in the supplement state, the negative control outcomes should not be known indicators of type 2 diabetes severity. We thus removed “essential (primary) hypertension” and “hyperlipidemia/hypercholesterolemia” from our list of negative control outcomes due to their known association with cardiovascular comorbidities, an index component of the Diabetes Complications and Severity Index. We replaced those two negative control outcomes with the two next-most prevalent outcomes satisfying our criteria: “syncope and collapse” and “unspecified asthma uncomplicated.”

### Supplementary Tables to Support Replication

**Table S5.** Cohort Criteria

| Cohort | ICD-10 Codes |
| --- | --- |
| Prediabetes | R73% |
| Type 2 Diabetes | E11% |

**Table S6.** Drug Class Members

| Medication Class | Generic Name |
| --- | --- |
| Biguanides (metformin) | alogliptin-metformin hcl, canagliflozin-metformin hcl, dapagliflozin-metformin hcl, empagliflozin-metformin hcl, ertugliflozin-metformin hcl, glipizide-metformin hcl, glyburide-metformin, linagliptin-metformin hcl, metformin hcl, pioglitazone hcl-metformin hcl, repaglinide-metformin hcl, rosiglitazone maleate-metformin hcl, saxagliptin-metformin hcl, sitagliptin-metformin hcl |
| Insulins | insulin aspart, insulin aspart (with niacinamide), insulin aspart protamine & aspart (human), insulin glulisine, insulin lispro, insulin lispro protamine & lispro, insulin lispro-aabc, insulin nph isophane & reg (human), insulin reg (human) buffered, insulin regular, insulin regular (human), insulin regular (human) in sodium chloride, insulin regular (pork), insulin degludec, insulin degludec-liraglutide, insulin detemir, insulin glargine, insulin glargine-lixisenatide, insulin isophane, insulin isophane (pork), insulin nph (human) (isophane), insulin zinc, insulin zinc (human), insulin zinc (pork), insulin zinc extended (human) |
| Other Diabetes Drugs | pioglitazone hcl/glimepiride, glyburide, chlorpropamide, glipizide, glimepiride, tolbutamide, tolazamide, pioglitazone hcl, rosiglitazone maleate, miglitol, acarbose, pramlintide acetate, bromocriptine mesylate, sitagliptin phosphate, linagliptin, alogliptin benzoate, saxagliptin hcl, ertugliflozin/sitagliptin, dapagliflozin/saxagliptin hcl, empagliflozin/linagliptin, alogliptin benz/pioglitazone, dulaglutide, exenatide microspheres, lixisenatide, exenatide, albiglutide, liraglutide, semaglutide, nateglinide, repaglinide, empagliflozin, canagliflozin, dapagliflozin propanediol, ertugliflozin pidolate |

**Table S7.** Covariate Logic

| Covariate | Logic |
| --- | --- |
| Influenza Vaccination | American Hospital Formulary Service (AHFS) therapeutic class code 80120000 and generic name containing "flu" (pharmacy claims)<br>-OR-any of the following procedure codes: 90630, 90653, 90656, 90662, 90673, 90674, 90682, 90685, 90686, 90687, 90688, 90756, Q2039, Q2035, Q2037 (medical claims) |
| Physician Visit / Wellness Visit | either of the following health care encounter service type descriptions: "physician visits" or "wellness visits" (medical claims) |
| Elixhauser In-hospital Mortality Score | The Elixhauser In-Hospital Mortality Score follows the guidelines presented by Moore et al.[18] |
| Diabetes Complications Severity Index (DCSI) | The DCSI component scores follow logic presented by Glasheen et al. (appendices A1-A7, B).[19] In addition to standard insurance claims data, the database used for the study also had the necessary lab results to compute the nephrology component score per the indicated reference. |

**Table S8.**
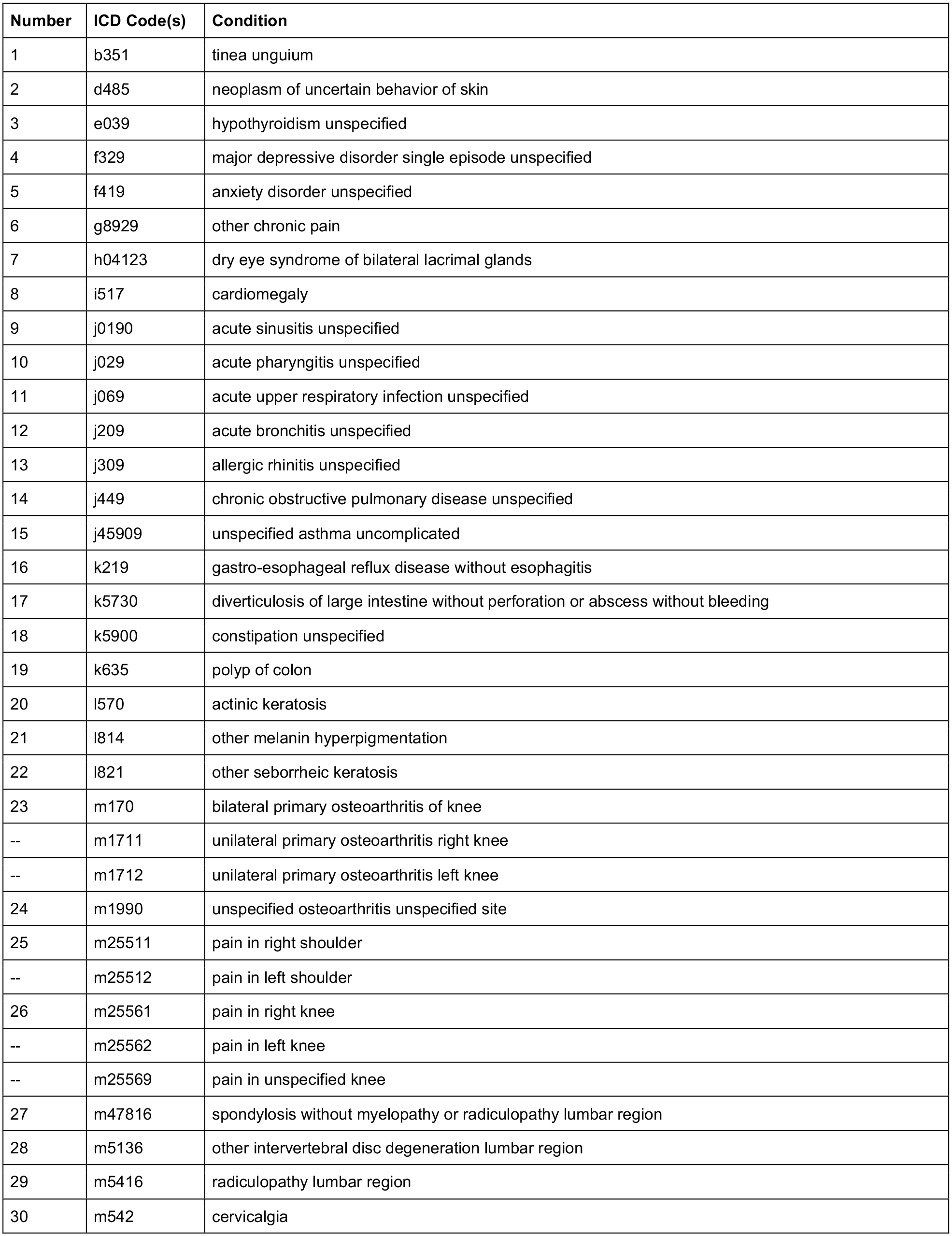
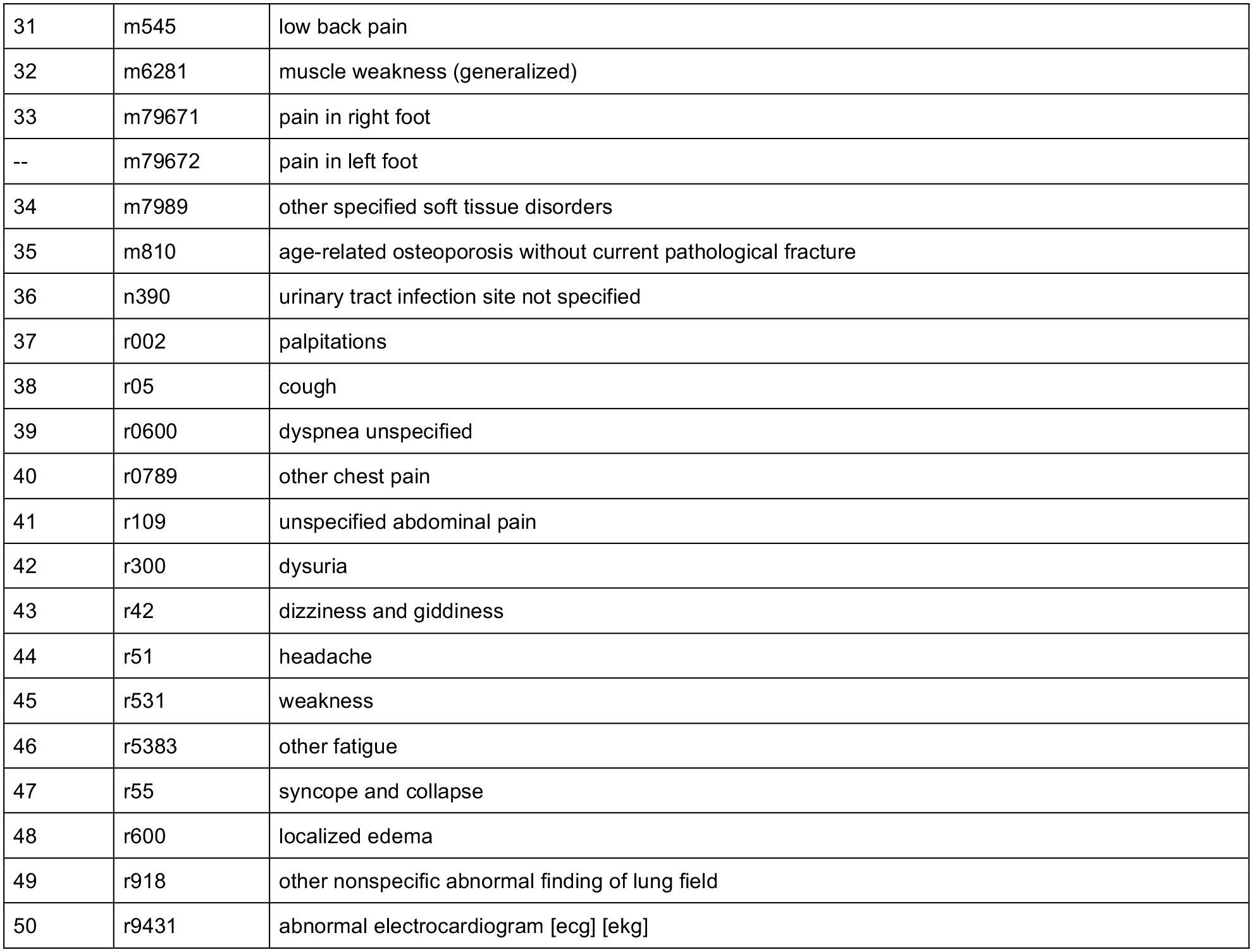
Negative Control Outcomes (50 total)

| Number | ICD Code(s) | Condition |
| --- | --- | --- |
| 1 | b351 | tinea unguium |
| 2 | d485 | neoplasm of uncertain behavior of skin |
| 3 | e039 | hypothyroidism unspecified |
| 4 | f329 | major depressive disorder single episode unspecified |
| 5 | f419 | anxiety disorder unspecified |
| 6 | g8929 | other chronic pain |
| 7 | h04123 | dry eye syndrome of bilateral lacrimal glands |
| 8 | i517 | cardiomegaly |
| 9 | j0190 | acute sinusitis unspecified |
| 10 | j029 | acute pharyngitis unspecified |
| 11 | j069 | acute upper respiratory infection unspecified |
| 12 | j209 | acute bronchitis unspecified |
| 13 | j309 | allergic rhinitis unspecified |
| 14 | j449 | chronic obstructive pulmonary disease unspecified |
| 15 | j45909 | unspecified asthma uncomplicated |
| 16 | k219 | gastro-esophageal reflux disease without esophagitis |
| 17 | k5730 | diverticulosis of large intestine without perforation or abscess without bleeding |
| 18 | k5900 | constipation unspecified |
| 19 | k635 | polyp of colon |
| 20 | l570 | actinic keratosis |
| 21 | l814 | other melanin hyperpigmentation |
| 22 | l821 | other seborrheic keratosis |
| 23 | m170 | bilateral primary osteoarthritis of knee |
| -- | m1711 | unilateral primary osteoarthritis right knee |
| -- | m1712 | unilateral primary osteoarthritis left knee |
| 24 | m1990 | unspecified osteoarthritis unspecified site |
| 25 | m25511 | pain in right shoulder |
| -- | m25512 | pain in left shoulder |
| 26 | m25561 | pain in right knee |
| -- | m25562 | pain in left knee |
| -- | m25569 | pain in unspecified knee |
| 27 | m47816 | spondylosis without myelopathy or radiculopathy lumbar region |
| 28 | m5136 | other intervertebral disc degeneration lumbar region |
| 29 | m5416 | radiculopathy lumbar region |
| 30 | m542 | cervicalgia |
| 31 | m545 | low back pain |
| 32 | m6281 | muscle weakness (generalized) |
| 33 | m79671 | pain in right foot |
| -- | m79672 | pain in left foot |
| 34 | m7989 | other specified soft tissue disorders |
| 35 | m810 | age-related osteoporosis without current pathological fracture |
| 36 | n390 | urinary tract infection site not specified |
| 37 | r002 | palpitations |
| 38 | r05 | cough |
| 39 | r0600 | dyspnea unspecified |
| 40 | r0789 | other chest pain |
| 41 | r109 | unspecified abdominal pain |
| 42 | r300 | dysuria |
| 43 | r42 | dizziness and giddiness |
| 44 | r51 | headache |
| 45 | r531 | weakness |
| 46 | r5383 | other fatigue |
| 47 | r55 | syncope and collapse |
| 48 | r600 | localized edema |
| 49 | r918 | other nonspecific abnormal finding of lung field |
| 50 | r9431 | abnormal electrocardiogram [ecg] [ekg] |

### STROBE Statement

#### Checklist of items that should be included in reports of cohort studies

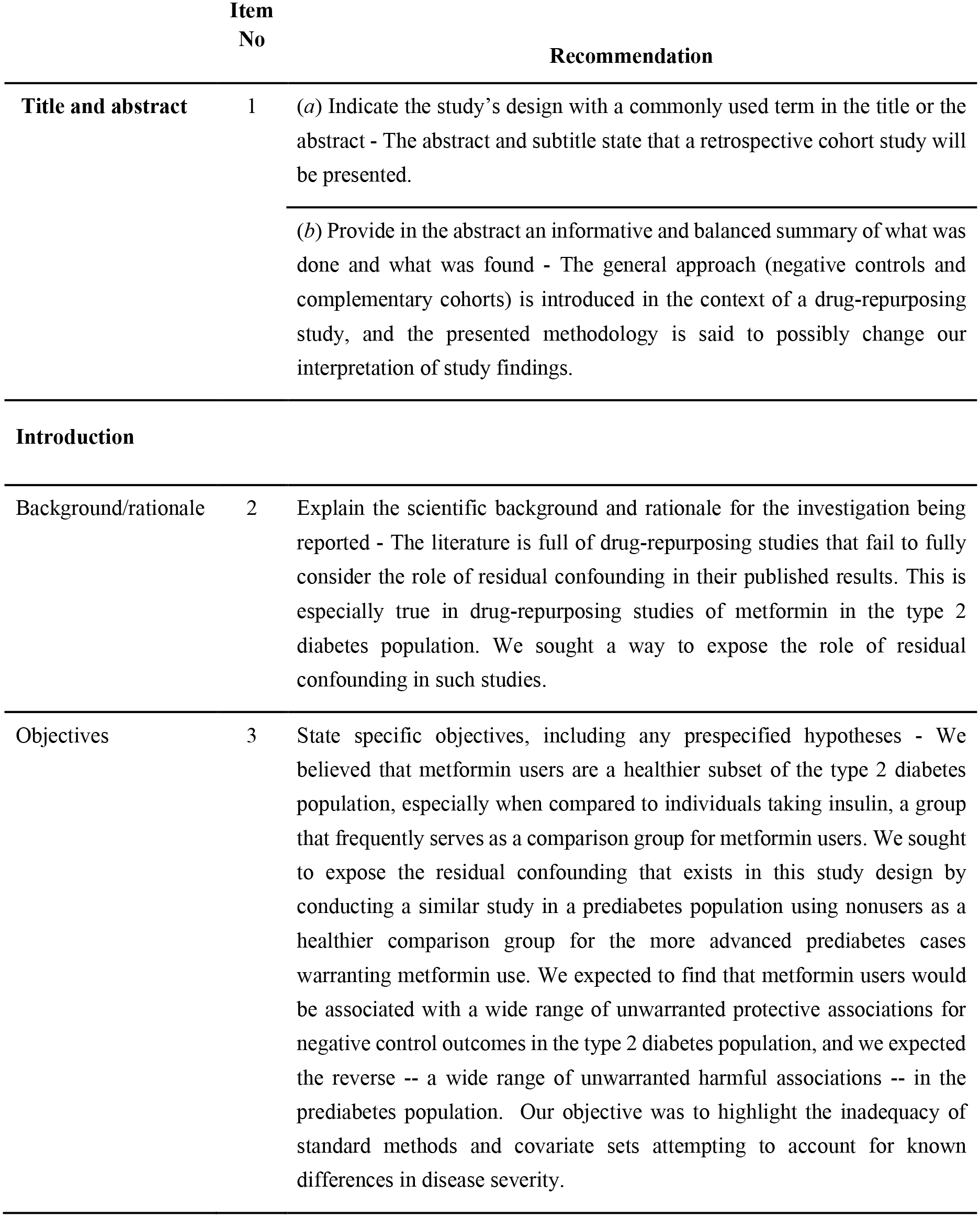
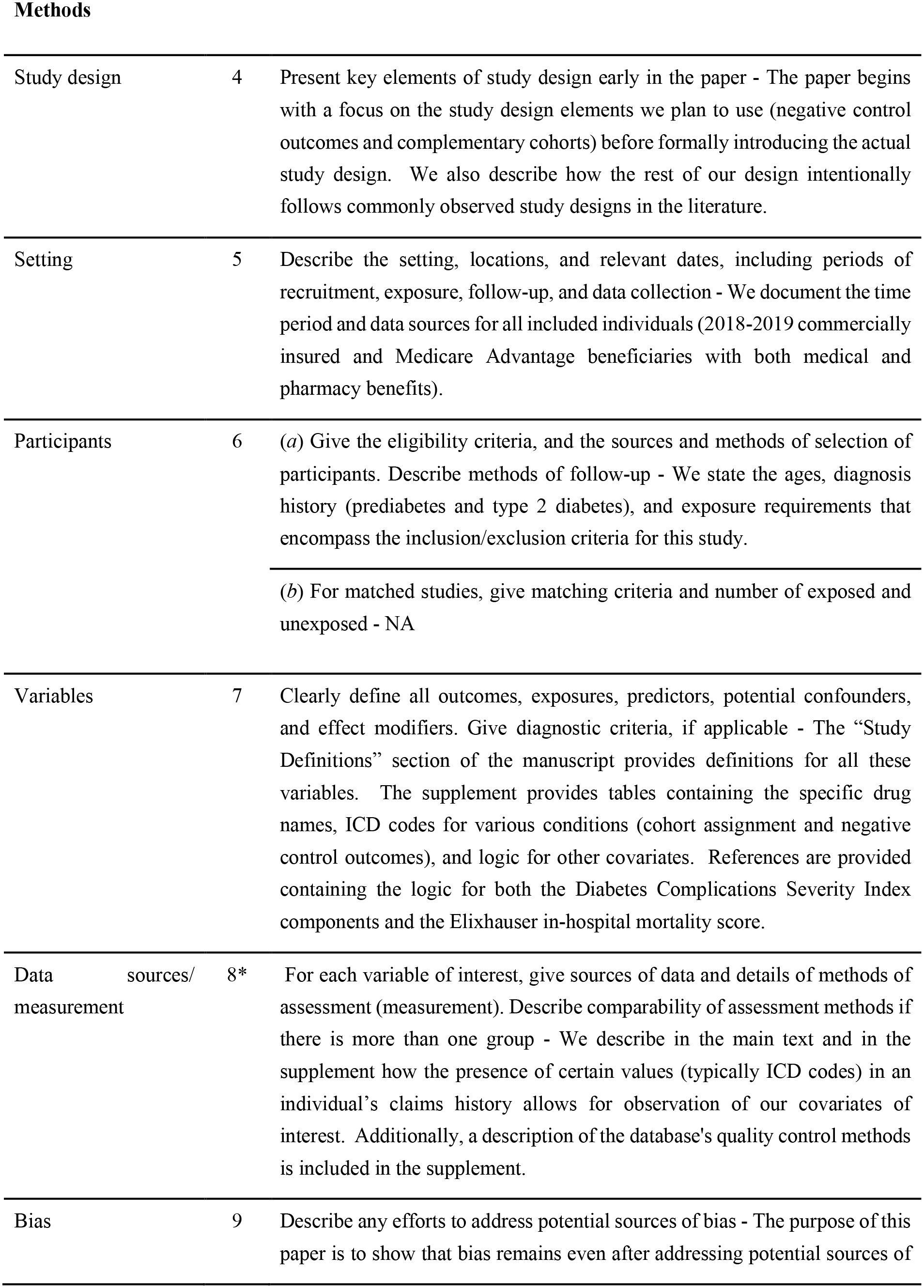
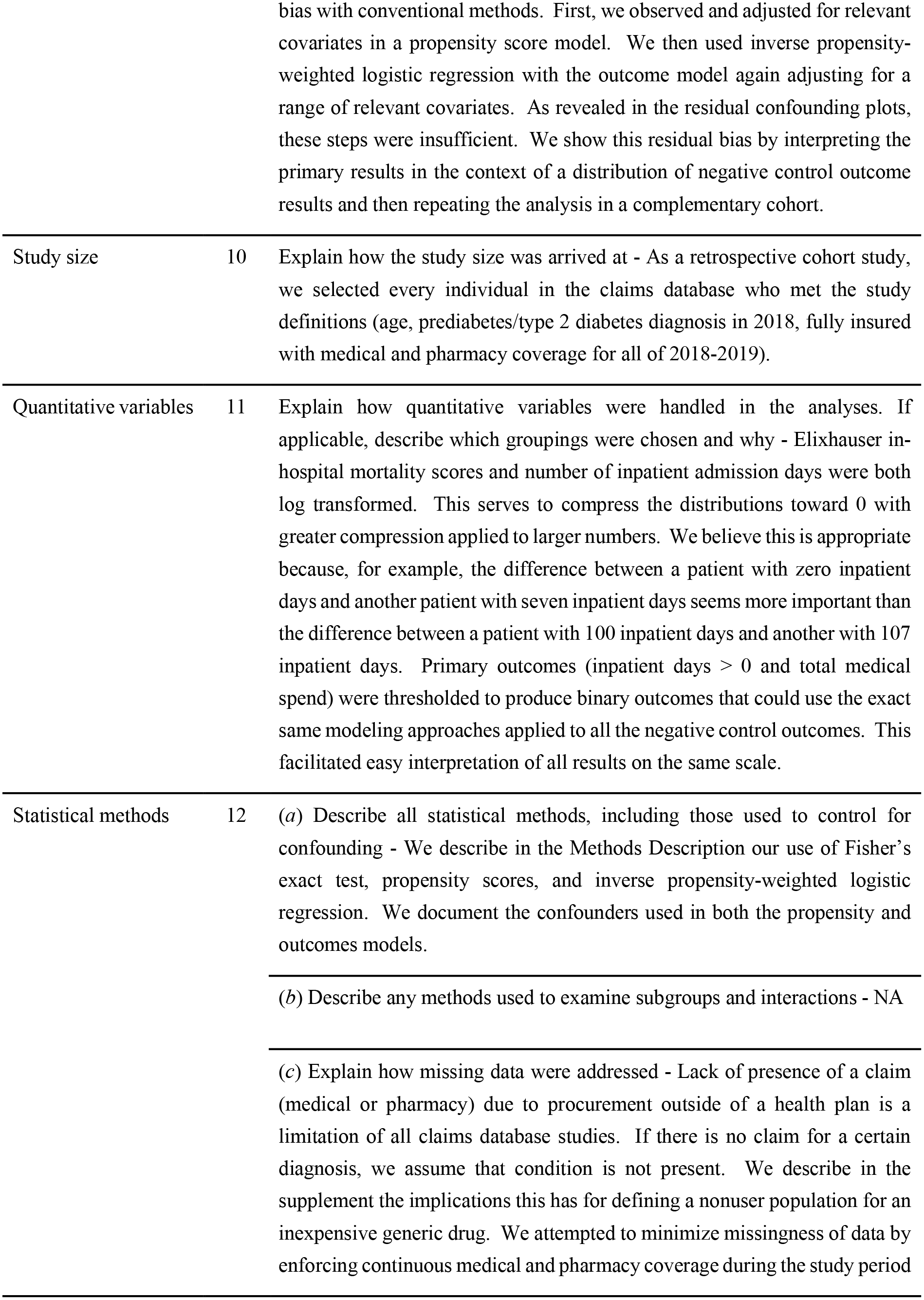
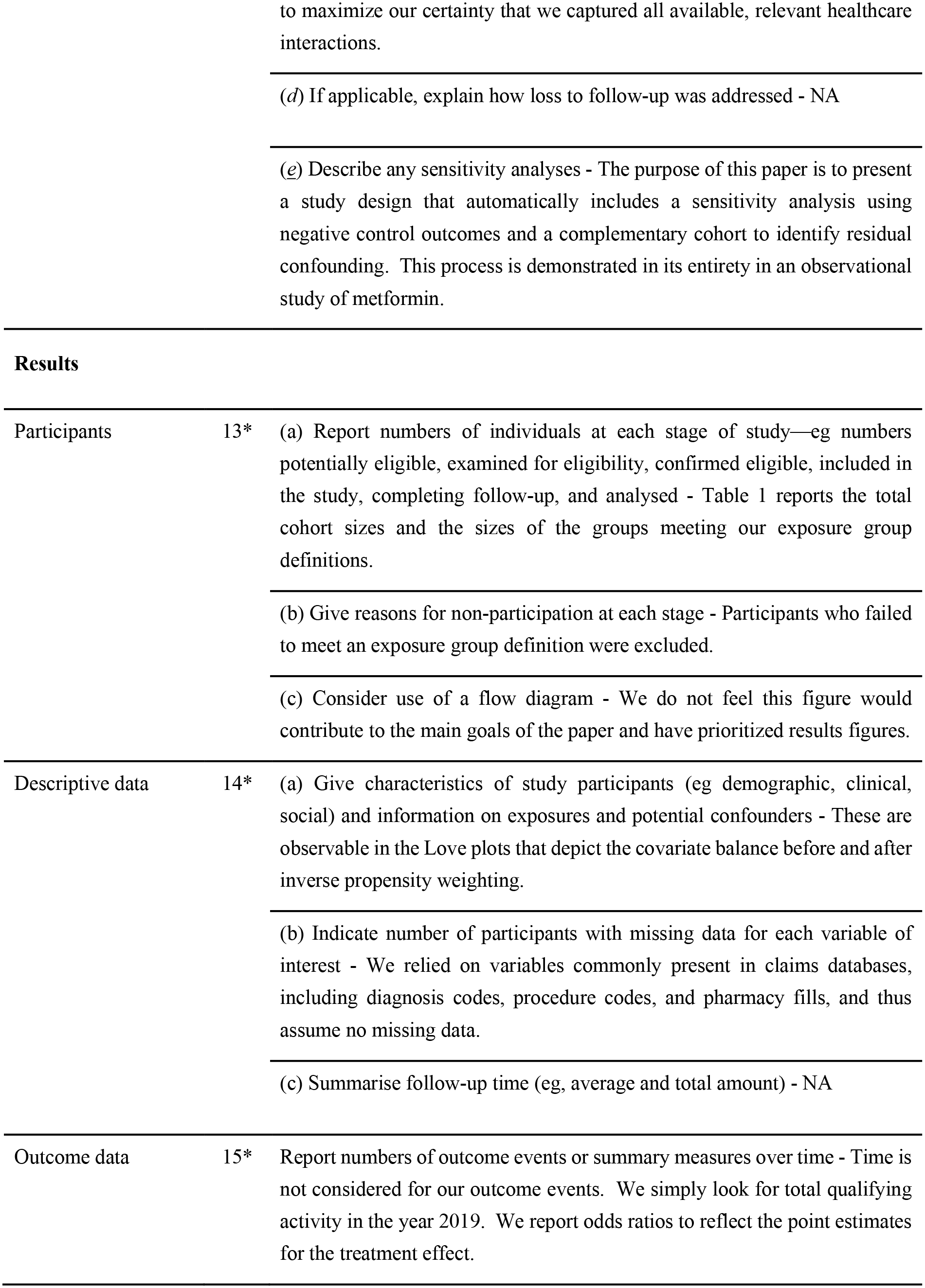
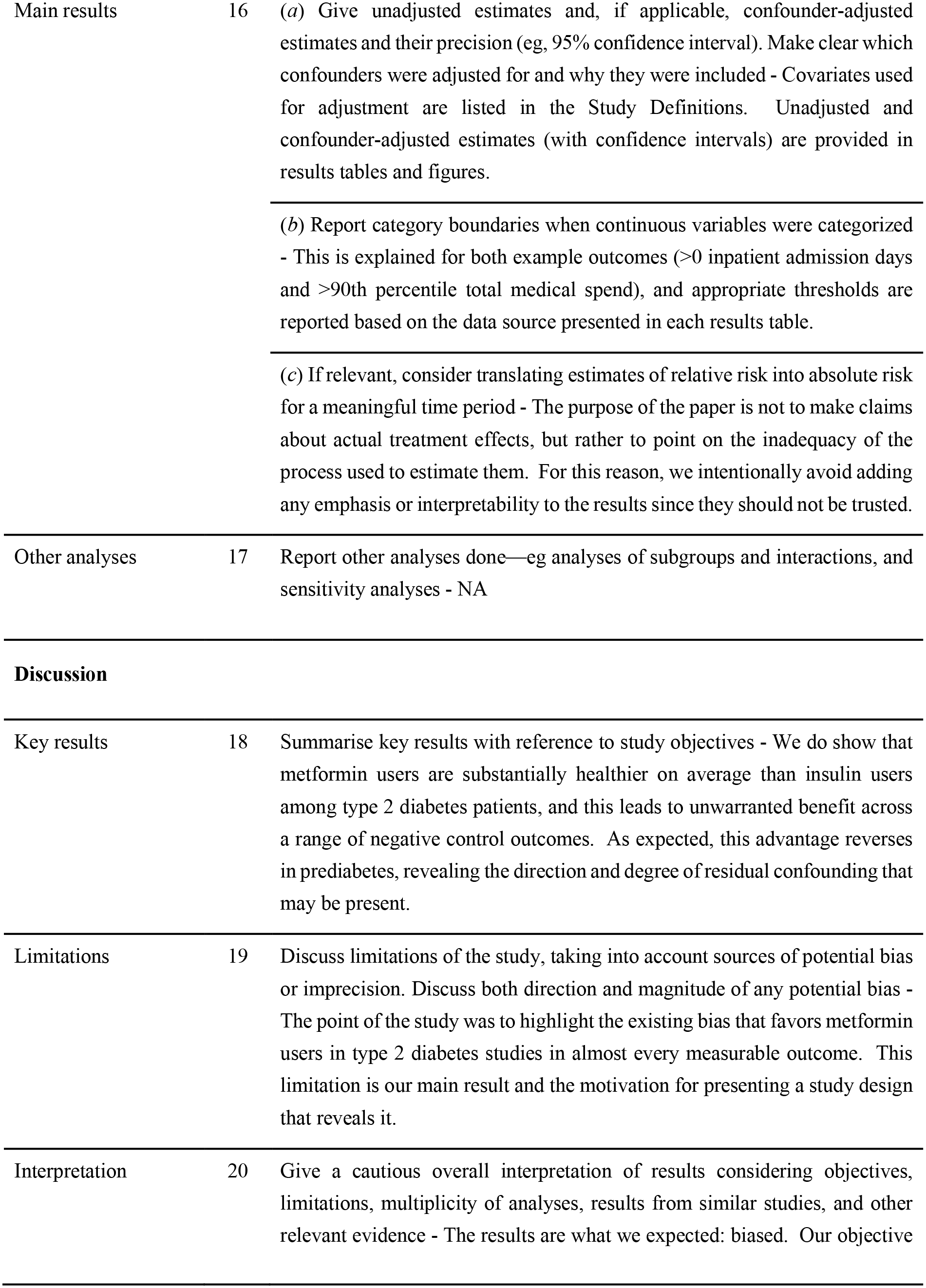
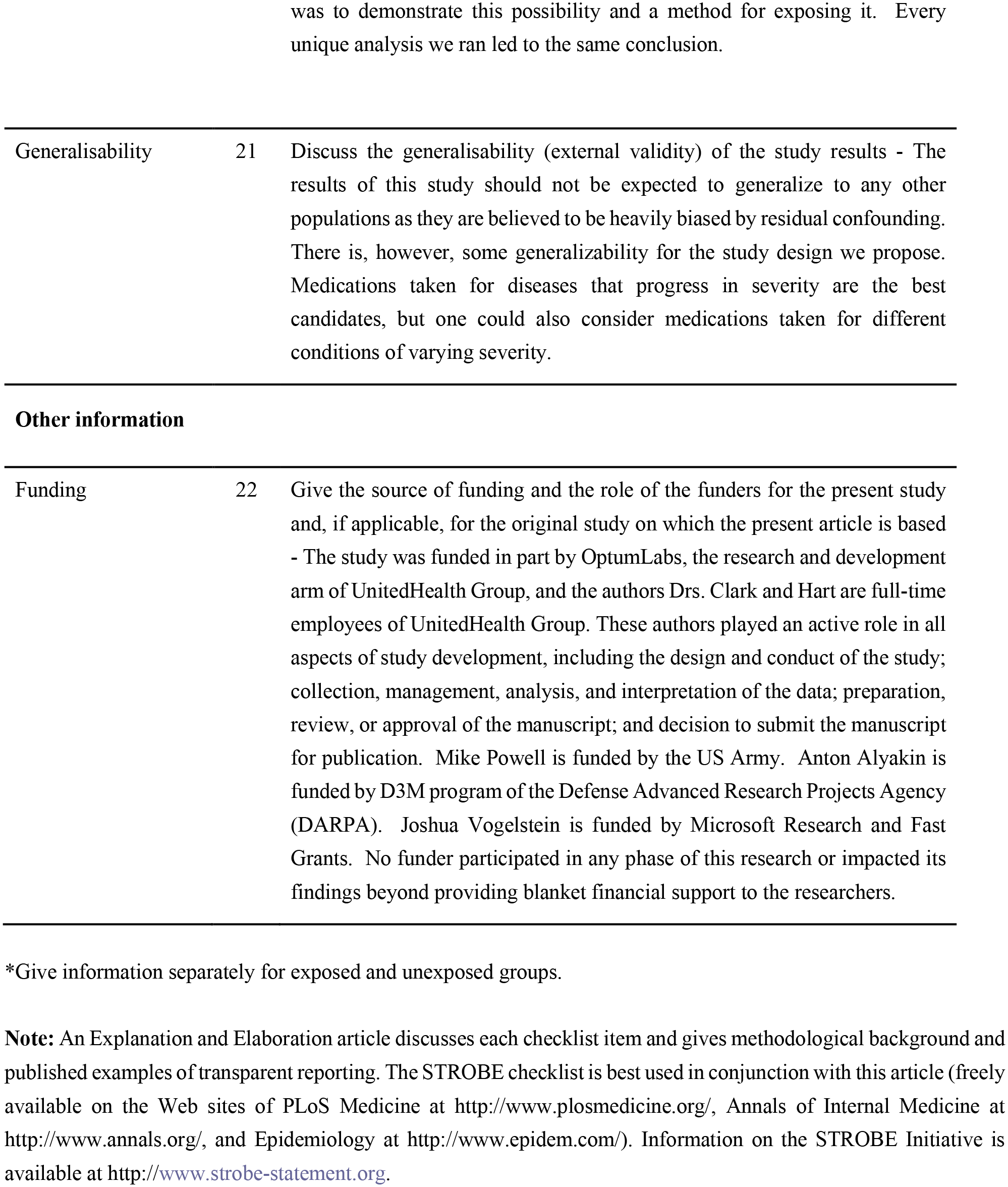

## Footnotes

1 The E-values represent the strength of association (on the risk ratio scale) an unobserved confounder must have with both the exposure and the outcome to nullify the effect estimates.

